# Adherence to a healthful plant-based diet and risk of chronic kidney disease among individuals with diabetes: A prospective cohort study

**DOI:** 10.1101/2024.03.14.24304283

**Authors:** Alysha S. Thompson, Anna Tresserra-Rimbau, Amy Jennings, Nicola P. Bondonno, Catharina J. Candussi, Joshua K. O’Neill, Claire Hill, Martina Gaggl, Aedín Cassidy, Tilman Kühn

**Author notes:** **Correspondence:** and **Corresponding Authors:** Prof Dr Tilman Kühn and Prof Aedín Cassidy Queen’s University Belfast Institute for Global Food Security (IGFS) / School of Biological Sciences 19 Chlorine Gardens Belfast BT9 5DL UK. **Funding Source:** Alysha S. Thompson holds a PhD studentship of the Department for the Economy (DfE), Northern Ireland.

## Abstract

**Background:** Chronic kidney disease (CKD) is highly prevalent among people with diabetes. While identifying modifiable risk factors to prevent a decline in kidney function among those living with diabetes is pivotal, there is limited evidence on dietary risk factors for CKD. In this study we examined the associations between healthy and less healthy plant-based diets (PBDs) and the risk of CKD among those with diabetes, and to identify potential underlying mechanisms.

**Methods:** We conducted a prospective analysis among 7,747 UK Biobank participants with prevalent diabetes. Multivariable Cox proportional hazard regression models were used to examine the associations between healthful and unhealthful PBDs and the risk of CKD. Causal mediation analyses were further employed to explore the underlying mechanisms of the observed associations.

**Results:** Among 7,747 study participants with diabetes, 1,030 developed incident CKD over 10.2 years of follow-up. Higher adherence to a healthy PBD was associated with a 24% lower CKD risk (HR_Q4 versus Q1_: 0.76 [95%CI: 0.63-0.92], p_trend_ = 0.002), while higher adherence to an unhealthy PBD was associated with a 35% higher risk (HR_Q4 versus Q1_: 1.35 [95%CI: 1.11-1.65], p_trend_ = 0.006). The observed associations were predominantly mediated by markers of body fatness (proportion mediated: 11-25%) and kidney function (23-89%).

**Conclusions:** In this prospective cohort study of middle-aged adults with diabetes, adherence to a healthy PBD was associated with lower CKD risk, whereas adherence to an unhealthy PBD was associated with a higher CKD risk. Associations were primarily mediated by markers of lower body fatness and improved kidney function.

## 1. Introduction

Chronic kidney disease (CKD) is a major global health issue affecting over one-tenth of the world’s population, ranking among one of the leading causes of death according to the Global Burden of Disease Study in 2017 (1). CKD is of particular concern among individuals living with diabetes, with 40% of new CKD cases being attributed to diabetes as the primary cause (1, 2). Although CKD is a comorbid condition causing progressive and irreversible loss of kidney function that bears a substantial economic burden worldwide (3, 4), differences in CKD risk among people with diabetes are not fully understood. Thus, identifying modifiable risk factors to aid in the preservation of kidney function and improve outcomes among those living with diabetes is pivotal for primary prevention.

Modifying dietary intake is a key component of diabetes management. It has been suggested that low-carbohydrate, Mediterranean, and Low-glycaemic Index dietary patterns are effective for improving outcomes for people with type 2 diabetes (5), while greater adherence to a Western diet (6), including higher intakes of ultra-processed foods (7, 8) has been associated with worse outcomes. The first study in the general population indicated that plant-based diets (PBD) are associated with lower CKD risk (9). This finding is in line with recent studies suggesting that replacing animal protein with plant protein is associated with lower CKD risk (10, 11). Conversely, a general reduction of protein intake is recommended as part of treatment guidelines for diabetic patients with CKD (12).

Given the preliminary evidence on a potential benefit of PBDs for CKD prevention, we hypothesized that a healthful PBD is related to lower CKD risk among people with diabetes. Therefore, the main objective of this study was to investigate whether PBDs, specifically adherence to a healthful vs unhealthful plant-based diet index (PDI), is associated with lower risks of CKD among individuals with diabetes, using data from the UK Biobank, a large-scale prospective cohort study. Our secondary objective was to assess which biological mechanisms may potentially mediate associations between PDI and CKD incidence among people with diabetes.

## 2. Methods

### 2.1 Study population

The UK Biobank is a large-scale population-based prospective cohort study of half a million adults aged between 40-69 years from the United Kingdom. The study recruited participants from 22 assessment centres across England, Scotland, and Wales between 2006 and 2010. At recruitment, study participants completed a baseline assessment comprising a touchscreen questionnaire, a face-to-face verbal interview, and a series of physical measurements.

Further details on the study protocol have been documented previously (13). The UK Biobank has received ethical approval from the NHS North West Multicentre Research Ethics Committee (Ref. 11/NW/0382). All participants provided signed informed consent at recruitment to participate in the study. We used the STROBE cohort checklist when writing this report (14).

Participants who had a prior diagnosis of diabetes and had completed at least one 24-hr dietary assessment were considered eligible for inclusion in this study. Participants were excluded if they withdrew their consent during the study follow-up, had implausible energy intakes (>17,573KJ or < 3,347KJ for men and > 14,644KJ or < 2,092KJ for women (15)), had prevalent CKD (self-reported or hospital inpatient record of CKD diagnosed before recruitment), an estimated glomerular filtration rate (eGFR) <60ml/min/1.73m^2^, were diagnosed with CKD before the final 24hr dietary assessment, or if follow-up ended before completion of the final dietary assessment (**Figure S1**).

### 2.2 Dietary assessment

Dietary information was collected using the Oxford WebQ 24-hour dietary assessment (16). The Oxford WebQ dietary questionnaire was issued on five separate occasions between April 2009 and June 2012, permitting up to 210,947 UK Biobank participants to complete at least one questionnaire. Mean (SD) duration (years) between baseline and first and last dietary assessments were 1.5 (1.4) and 2.2 (1.4), respectively. The Oxford WebQ has been validated to represent a true approximation of dietary intake through comparison to objective nutritional biomarkers and an interviewer-administrated 24-hr dietary recall (17–19).

### 2.3 Plant-based diet indices

Following methods described in previous studies (20–22), the Oxford WebQ dietary questionnaire (16, 23) was used to calculate a healthful plant-based diet index (hPDI) and an unhealthful plant-based diet index (uPDI). A total of 17 food groups (whole grains, fruits, vegetables, nuts, legumes and vegetarian protein alternatives, tea and coffee, fruit juices, refined grains, potatoes, sugar-sweetened beverages (SSBs), sweets and desserts, animal fat, dairy, eggs, fish or seafood, meat, and miscellaneous animal-based foods) were used to calculate the PDIs. Although used previously as a component of the PDI, data on vegetable oils was not available for use in this study (22). The 17 food groups were further classified into a hPDI and a uPDI. The hPDI consisted of scoring healthy plant-food groups (whole grains, fruits, vegetables, nuts, legumes and vegetarian protein alternatives and tea and coffee) positively, and unhealthy plant-food groups (fruit juices, refined grains, potatoes, SSBs, sweets and desserts) and animal-based food groups (animal fat, dairy, eggs, fish or seafood, meat, and miscellaneous animal-based foods) negatively. For the uPDI, unhealthy plant-food groups were scored positively, while healthy plant-food groups and animal-based food groups were scored negatively. Participants with intakes of more than zero portions were ranked into quartiles of intake for each of the 17 food groups. For positive scores, participants were assigned a score between 2 and 5 (2 for the lowest quartile of intake and 5 for the highest) for each food group. Those with intakes of zero were assigned a score of 1. For reverse scores, the scoring pattern was inverted. To calculate hPDI and uPDI, the scores assigned to each participant across the 17 food groups were summed and categorised into sex-specific quartiles.

### 2.4 Ascertainment of diabetes

Prevalent diabetes was defined using the algorithms reported previously (24). Participants were considered as having prevalent diabetes if they were diagnosed with diabetes (self-reported or hospital inpatient diagnosed) before completing the first dietary assessment, reported taking anti-diabetic medications, had a random blood glucose ≥11.1 mmol/l, or had a haemoglobin A1c (HBA1c) level of ≥6.5%.

### 2.5 Ascertainment of CKD

The primary outcome of this study was incident CKD. Incident CKD cases were defined using the International Classification of Diseases, 10^th^ Revision (ICD-10) codes in Hospital Inpatient data and Death Registry data (ICD-10 codes: N03, N06, N08, N11, N12, N13, N14, N15, N16, N18, N19, Z49, I12, I13) or Office of Population Censuses and Surveys Classification of Interventions and Procedures-version 4 (OPCS-4) codes in Hospital Inpatient data (OPCS-4 codes: L74.1-L74.6, L74.8, L74.9, M01.2, M01.3-M01.5, M01.8, M01.9, M02.3, M08.4, M17.2, M17.4, M17.8, M17.9, X40.1-X40.9, X41.1, X41.2, X41.8, X41.9, X42.1, X42.8, X42.9, and X43.1). Follow-up time was calculated from the date of the last dietary assessment and was censored at the date of CKD diagnosis, death, lost to follow-up or end of follow-up, whichever occurred first. Hospital admission data was available up until 31^st^ October 2022 from the Hospital Episode Statistics (HES) for participants in England, 31^st^ August 2022 from the Scottish Morbidity Records (SMR) for participants in Scotland and 31^st^ May 2022 from the Patient Episode Database for Wales (PEDW) for participants in Wales.

### 2.6 Assessment of covariates

Information on sociodemographic, dietary, lifestyle and medical history was collected at baseline via a touchscreen questionnaire and verbal interview between 2006 and 2010. Further, study participants had physical measurements and biological samples collected by a trained member of staff (13). Serum creatinine, measured by enzymatic analysis on a Beckman Coulter AU5800, was used to calculate eGFR using the CKD-EPI (Chronic Kidney Disease Epidemiology Collaboration) creatinine equation (25). Further details on data collection and classification of covariates used in this study can be found in **Supplementary Table S1.**

### 2.7 Assessment of genetic risk score of eGFR

The polygenic risk score (PRS) of eGFR was calculated by a weighted method using 161 single nucleotide polymorphisms (SNPs) of eGFR (**Supplementary Table S2**) identified by Wuttke et al. 2019. A higher eGFR PRS was indicative of a lower genetic predisposition to kidney diseases. Further information on the genotyping and quality control (QC) for this study can be found in the **Supplementary Methods**.

### 2.8 Statistical analysis

Baseline characteristics are presented as mean ± standard deviation (SD) for continuous variables and as proportions for categorical variables across quartiles of PBD scores. Cox proportional hazards regression models were used to examine the associations between PDIs and risk of CKD, producing hazard ratios (HR) and 95% confidence intervals (CIs). Linear trend tests were carried out by modelling the PDIs as continuous exposure variables (P-trend). Model 1 was adjusted for sex (female, male) and education (Low: CSEs or equivalent, O levels/GCSEs or equivalent; Medium: A levels/AS levels or equivalent, NVQ or HND or HNC or equivalent; High: College or University degree, other professional qualifications eg: nursing, teaching; unknown/missing/prefer not to say (15.1%)), stratified by age (<55 years, 55-<65, ≥65 years) and region (10 regions). Model 2 was further adjusted for body mass index (BMI) (18·5-24·9 kg/m^2^, 25·0-29·9 kg/m^2^, ≥30 kg/m^2^, or unknown/missing (0.3%)), waist circumference (continuous scale, cm), ethnicity (Asian, Black, Multiple, White, other/unknown/missing (1.8%)), physical activity (METs hr/week in quintiles, or unknown/missing (2.7%)), smoking status (never, previous, current, or unknown/missing (0.5%)), alcohol intake (continuous scale, g/day), energy intake (continuous scale, kJ/day), multimorbidity index (number of pre-existing long-term conditions; 0, 1, 2, or >3), Townsend deprivation index (quintiles from low to high deprivation index), type of diabetes (type 1, type 2, unspecified), diabetes medication use (no, yes, or missing (25.0%)) and number of completed dietary assessments (continuous scale, ranging between 1-5). Model 3 further included total protein intake (continuous scale, g/day) as a potential confounder.

Subgroup analyses were carried out to examine effect modification. Potential effect modifiers included, age (<60 or ≥60 years), sex (male or female), BMI (<25 or ≥25kg/m^2^), smoking status (never or ever), alcohol intake (low [<1g/day] or moderate [1-≤15g/day] or high [≥16g/day]), total protein intake (<median [<81g/day] or ≥ median [≥81g/day]), history of hypertension (no or yes), and diabetes type (type 1 or type 2). To assess the robustness of findings, several sensitivity analyses were carried out. Firstly, eGFR, nephrolithiasis and blood biomarkers, including HDL-cholesterol (HDL-C), LDL-cholesterol (LDL-C), lipoprotein(a), c-reactive protein (CRP) and albumin were further adjusted for. Secondly, to assess whether associations between adherence to the PDI and CKD were influenced by genetic susceptibility to kidney diseases, a PRS of CKD (eGFR) was further adjusted for.

Thirdly, primary analyses were repeated excluding participants with less than two years of follow-up time to account for potential reverse causality. Fourthly, primary analyses were repeated excluding sugar-sweetened beverages from the score, as these food items were most strongly associated with CKD risk on the food group level (see Results). Finally, the E-value methodology (27) was used to assess the overall strength of potential unmeasured confounding.

Mediation analyses were carried out to assess the mediating role of measures commonly associated with renal function on the PDI-CKD relationship. To conduct this mediation analysis, the *paramed* package in Stata (28, 29) was used. This approach uses parametric regression to estimate two models: a model for the mediator conditional on the exposure and covariates, and a model for the outcome conditional on the exposure, the mediator and covariates. Based on previous studies, the following mediators were chosen: markers of obesity and lipid metabolism (BMI (30), waist circumference (31), cholesterol (LDL-C, including lipoprotein A and apolipoprotein A, as well as HDL-C and triglycerides)) (30, 32), kidney function (eGFR (33), cystatin C (34), urate (35), creatinine (36) nephrolithiasis (37)), hypertension (30) and inflammation (CRP) (38). To calculate the percentage proportion of the association mediated through each potential mediator, the log of the natural indirect effect (NIE) HR was divided by the sum of the log of the natural indirect effect (NIE) HR and the log of the controlled direct effect (CDE) HR (log(NIE)/log(NIE)+log(NDE)).

A two-sided *P* <0.05 was considered statistically significant. All statistical analyses were carried out using Stata 18.0 (StataCorp LLC). Upon completing the Schoenfeld residuals test, there was no indication of violation of the proportional hazards assumption.

## 3. Results

### 3.1 Characteristics of the study population

Baseline characteristics, including key nutrient intakes of study participants are shown across quartiles of hPDI and uPDI in **Table 1 and Tables S3-S5 in the Supplement**. Of the 502,236 UK Biobank participants enrolled, 42,713 had prevalent diabetes, 8,308 of which had data available from 1 or more dietary assessments (**Figure S1**). In the current study, 7,747 (1.5%) participants had prevalent diabetes, 3,038 (39.2%) were female and 4,709 were male (60.8%). The mean (SD) age was 58.8 (7.2) years, 6,910 (89.2%) were White, and 3,829 (49.4%) had a BMI over ≥30kg/m^2^. Participants in the highest quartile of hPDI were older, were more likely to have a lower BMI, be more physically active, have a higher level of education, and have never smoked compared to participants in the lowest quartile.

**Table 1.**
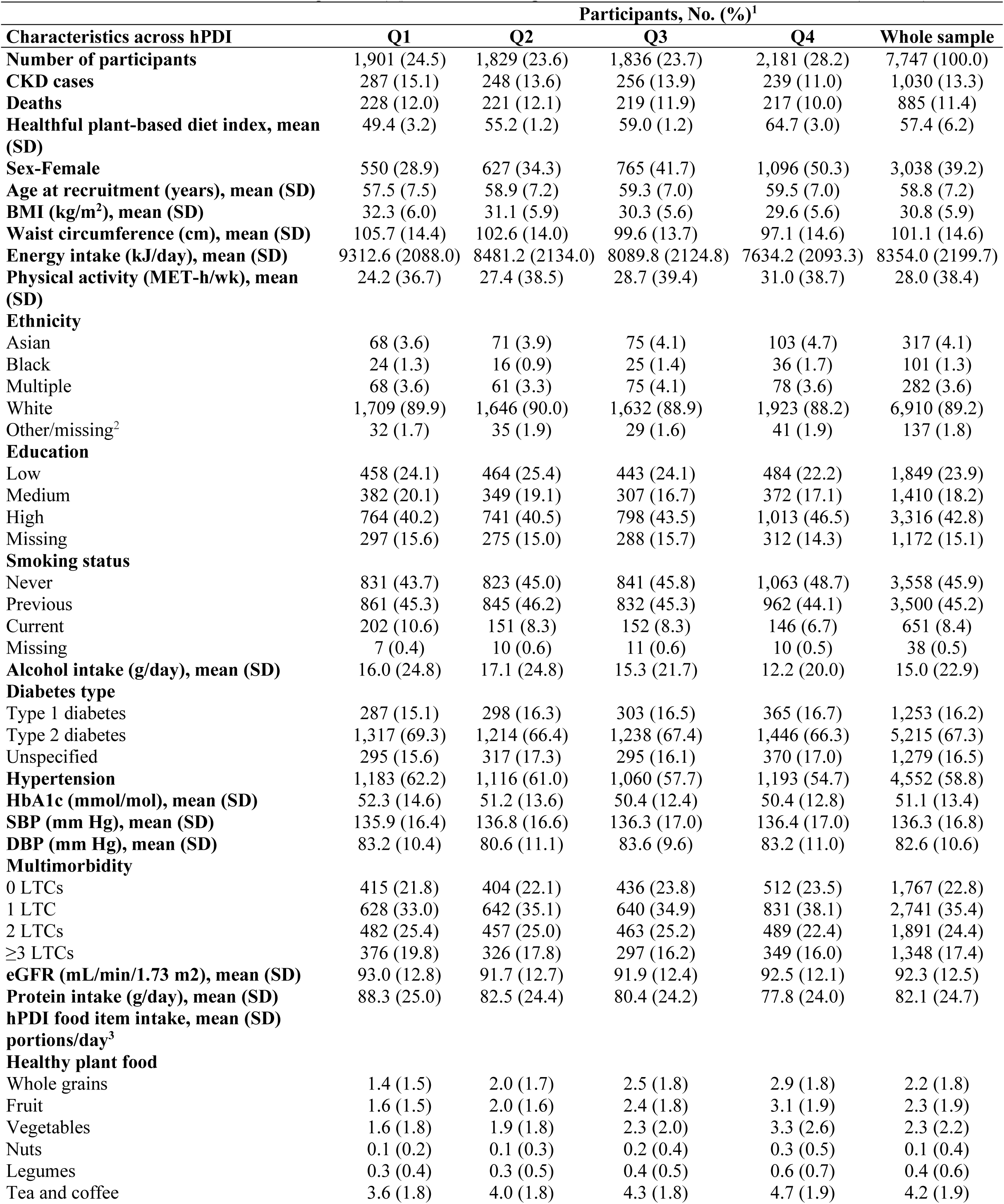
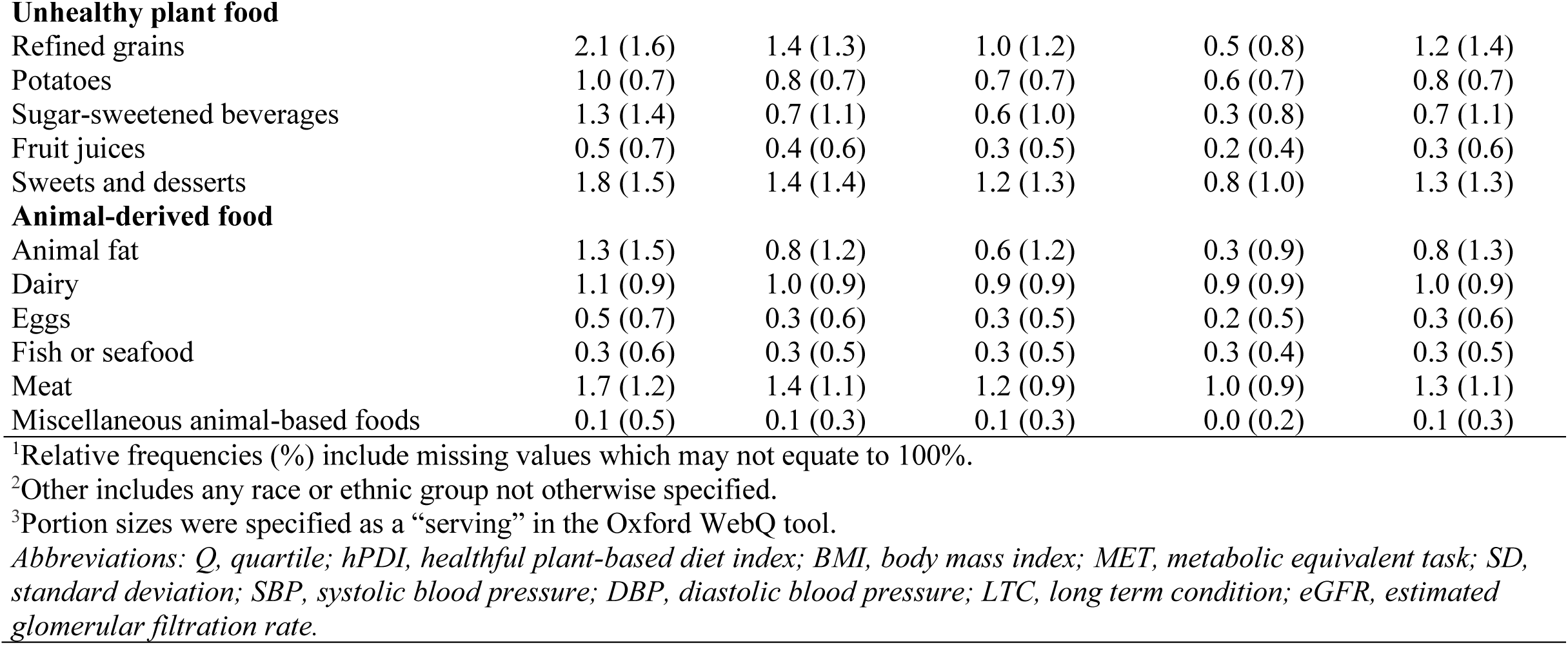
Baseline characteristics across quartiles (Q) of the healthful plant-based diet index in the UK Biobank (n=7,747)

| Characteristics across hPDI | Participants, No. (%)<sup>1</sup> |  |  |  |  |
| --- | --- | --- | --- | --- | --- |
|  | Q1 | Q2 | Q3 | Q4 | Whole sample |
| **Number of participants** | 1,901 (24.5) | 1,829 (23.6) | 1,836 (23.7) | 2,181 (28.2) | 7,747 (100.0) |
| **CKD cases** | 287 (15.1) | 248 (13.6) | 256 (13.9) | 239 (11.0) | 1,030 (13.3) |
| **Deaths** | 228 (12.0) | 221 (12.1) | 219 (11.9) | 217 (10.0) | 885 (11.4) |
| **Healthful plant-based diet index, mean (SD)** | 49.4 (3.2) | 55.2 (1.2) | 59.0 (1.2) | 64.7 (3.0) | 57.4 (6.2) |
| **Sex-Female** | 550 (28.9) | 627 (34.3) | 765 (41.7) | 1,096 (50.3) | 3,038 (39.2) |
| **Age at recruitment (years), mean (SD)** | 57.5 (7.5) | 58.9 (7.2) | 59.3 (7.0) | 59.5 (7.0) | 58.8 (7.2) |
| **BMI (kg/m<sup>2</sup>), mean (SD)** | 32.3 (6.0) | 31.1 (5.9) | 30.3 (5.6) | 29.6 (5.6) | 30.8 (5.9) |
| **Waist circumference (cm), mean (SD)** | 105.7 (14.4) | 102.6 (14.0) | 99.6 (13.7) | 97.1 (14.6) | 101.1 (14.6) |
| **Energy intake (kJ/day), mean (SD)** | 9312.6 (2088.0) | 8481.2 (2134.0) | 8089.8 (2124.8) | 7634.2 (2093.3) | 8354.0 (2199.7) |
| **Physical activity (MET-h/wk), mean (SD)** | 24.2 (36.7) | 27.4 (38.5) | 28.7 (39.4) | 31.0 (38.7) | 28.0 (38.4) |
| **Ethnicity** |  |  |  |  |  |
| Asian | 68 (3.6) | 71 (3.9) | 75 (4.1) | 103 (4.7) | 317 (4.1) |
| Black | 24 (1.3) | 16 (0.9) | 25 (1.4) | 36 (1.7) | 101 (1.3) |
| Multiple | 68 (3.6) | 61 (3.3) | 75 (4.1) | 78 (3.6) | 282 (3.6) |
| White | 1,709 (89.9) | 1,646 (90.0) | 1,632 (88.9) | 1,923 (88.2) | 6,910 (89.2) |
| Other/missing<sup>2</sup> | 32 (1.7) | 35 (1.9) | 29 (1.6) | 41 (1.9) | 137 (1.8) |
| **Education** |  |  |  |  |  |
| Low | 458 (24.1) | 464 (25.4) | 443 (24.1) | 484 (22.2) | 1,849 (23.9) |
| Medium | 382 (20.1) | 349 (19.1) | 307 (16.7) | 372 (17.1) | 1,410 (18.2) |
| High | 764 (40.2) | 741 (40.5) | 798 (43.5) | 1,013 (46.5) | 3,316 (42.8) |
| Missing | 297 (15.6) | 275 (15.0) | 288 (15.7) | 312 (14.3) | 1,172 (15.1) |
| **Smoking status** |  |  |  |  |  |
| Never | 831 (43.7) | 823 (45.0) | 841 (45.8) | 1,063 (48.7) | 3,558 (45.9) |
| Previous | 861 (45.3) | 845 (46.2) | 832 (45.3) | 962 (44.1) | 3,500 (45.2) |
| Current | 202 (10.6) | 151 (8.3) | 152 (8.3) | 146 (6.7) | 651 (8.4) |
| Missing | 7 (0.4) | 10 (0.6) | 11 (0.6) | 10 (0.5) | 38 (0.5) |
| **Alcohol intake (g/day), mean (SD)** | 16.0 (24.8) | 17.1 (24.8) | 15.3 (21.7) | 12.2 (20.0) | 15.0 (22.9) |
| **Diabetes type** |  |  |  |  |  |
| Type 1 diabetes | 287 (15.1) | 298 (16.3) | 303 (16.5) | 365 (16.7) | 1,253 (16.2) |
| Type 2 diabetes | 1,317 (69.3) | 1,214 (66.4) | 1,238 (67.4) | 1,446 (66.3) | 5,215 (67.3) |
| Unspecified | 295 (15.6) | 317 (17.3) | 295 (16.1) | 370 (17.0) | 1,279 (16.5) |
| **Hypertension** | 1,183 (62.2) | 1,116 (61.0) | 1,060 (57.7) | 1,193 (54.7) | 4,552 (58.8) |
| **HbA1c (mmol/mol), mean (SD)** | 52.3 (14.6) | 51.2 (13.6) | 50.4 (12.4) | 50.4 (12.8) | 51.1 (13.4) |
| **SBP (mm Hg), mean (SD)** | 135.9 (16.4) | 136.8 (16.6) | 136.3 (17.0) | 136.4 (17.0) | 136.3 (16.8) |
| **DBP (mm Hg), mean (SD)** | 83.2 (10.4) | 80.6 (11.1) | 83.6 (9.6) | 83.2 (11.0) | 82.6 (10.6) |
| **Multimorbidity** |  |  |  |  |  |
| 0 LTCs | 415 (21.8) | 404 (22.1) | 436 (23.8) | 512 (23.5) | 1,767 (22.8) |
| 1 LTC | 628 (33.0) | 642 (35.1) | 640 (34.9) | 831 (38.1) | 2,741 (35.4) |
| 2 LTCs | 482 (25.4) | 457 (25.0) | 463 (25.2) | 489 (22.4) | 1,891 (24.4) |
| ≥3 LTCs | 376 (19.8) | 326 (17.8) | 297 (16.2) | 349 (16.0) | 1,348 (17.4) |
| **eGFR (mL/min/1.73 m<sup>2</sup>), mean (SD)** | 93.0 (12.8) | 91.7 (12.7) | 91.9 (12.4) | 92.5 (12.1) | 92.3 (12.5) |
| **Protein intake (g/day), mean (SD)** | 88.3 (25.0) | 82.5 (24.4) | 80.4 (24.2) | 77.8 (24.0) | 82.1 (24.7) |
| **hPDI food item intake, mean (SD) portions/day<sup>3</sup>** |  |  |  |  |  |
| **Healthy plant food** |  |  |  |  |  |
| Whole grains | 1.4 (1.5) | 2.0 (1.7) | 2.5 (1.8) | 2.9 (1.8) | 2.2 (1.8) |
| Fruit | 1.6 (1.5) | 2.0 (1.6) | 2.4 (1.8) | 3.1 (1.9) | 2.3 (1.9) |
| Vegetables | 1.6 (1.8) | 1.9 (1.8) | 2.3 (2.0) | 3.3 (2.6) | 2.3 (2.2) |
| Nuts | 0.1 (0.2) | 0.1 (0.3) | 0.2 (0.4) | 0.3 (0.5) | 0.1 (0.4) |
| Legumes | 0.3 (0.4) | 0.3 (0.5) | 0.4 (0.5) | 0.6 (0.7) | 0.4 (0.6) |
| Tea and coffee | 3.6 (1.8) | 4.0 (1.8) | 4.3 (1.8) | 4.7 (1.9) | 4.2 (1.9) |
| **Unhealthy plant food** |  |  |  |  |  |
| Refined grains | 2.1 (1.6) | 1.4 (1.3) | 1.0 (1.2) | 0.5 (0.8) | 1.2 (1.4) |
| Potatoes | 1.0 (0.7) | 0.8 (0.7) | 0.7 (0.7) | 0.6 (0.7) | 0.8 (0.7) |
| Sugar-sweetened beverages | 1.3 (1.4) | 0.7 (1.1) | 0.6 (1.0) | 0.3 (0.8) | 0.7 (1.1) |
| Fruit juices | 0.5 (0.7) | 0.4 (0.6) | 0.3 (0.5) | 0.2 (0.4) | 0.3 (0.6) |
| Sweets and desserts | 1.8 (1.5) | 1.4 (1.4) | 1.2 (1.3) | 0.8 (1.0) | 1.3 (1.3) |
| **Animal-derived food** |  |  |  |  |  |
| Animal fat | 1.3 (1.5) | 0.8 (1.2) | 0.6 (1.2) | 0.3 (0.9) | 0.8 (1.3) |
| Dairy | 1.1 (0.9) | 1.0 (0.9) | 0.9 (0.9) | 0.9 (0.9) | 1.0 (0.9) |
| Eggs | 0.5 (0.7) | 0.3 (0.6) | 0.3 (0.5) | 0.2 (0.5) | 0.3 (0.6) |
| Fish or seafood | 0.3 (0.6) | 0.3 (0.5) | 0.3 (0.5) | 0.3 (0.4) | 0.3 (0.5) |
| Meat | 1.7 (1.2) | 1.4 (1.1) | 1.2 (0.9) | 1.0 (0.9) | 1.3 (1.1) |
| Miscellaneous animal-based foods | 0.1 (0.5) | 0.1 (0.3) | 0.1 (0.3) | 0.0 (0.2) | 0.1 (0.3) |

<sup>1</sup>Relative frequencies (%) include missing values which may not equate to 100%.
<sup>2</sup>Other includes any race or ethnic group not otherwise specified.
<sup>3</sup>Portion sizes were specified as a “serving” in the Oxford WebQ tool.
*Abbreviations: Q, quartile; hPDI, healthful plant-based diet index; BMI, body mass index; MET, metabolic equivalent task; SD, standard deviation; SBP, systolic blood pressure; DBP, diastolic blood pressure; LTC, long term condition; eGFR, estimated glomerular filtration rate.*

Participants in the highest quartile of uPDI were younger, were more likely to have a higher BMI, be less physically active, have a lower level of education and have never smoked compared to participants in the lowest quartile.

### 3.2 Plant-based diets and chronic kidney disease risk

During 10.2 years of follow-up, 1,030 (13.3%) participants developed CKD. Minimal and multivariable-adjusted models, including a tiered modelling approach are presented in **Table 2**. In model 2, those in the highest quartile (Quartile 4 (Q4)) of hPDI had a 25% lower risk of CKD (HR: 0.75 [95%CI: 0.62-0.91], p_trend_ = 0.001) compared to those in the lowest quartile of hPDI (Quartile 1 (Q1)). Upon adjusting for total protein intake as a potential confounder in model 3, associations were only marginally attenuated with higher hPDI scores (Q4) being associated with a 24% lower risk of CKD (HR: 0.76 [95%CI: 0.63-0.92], p_trend_ = 0.002).

**Table 2.**
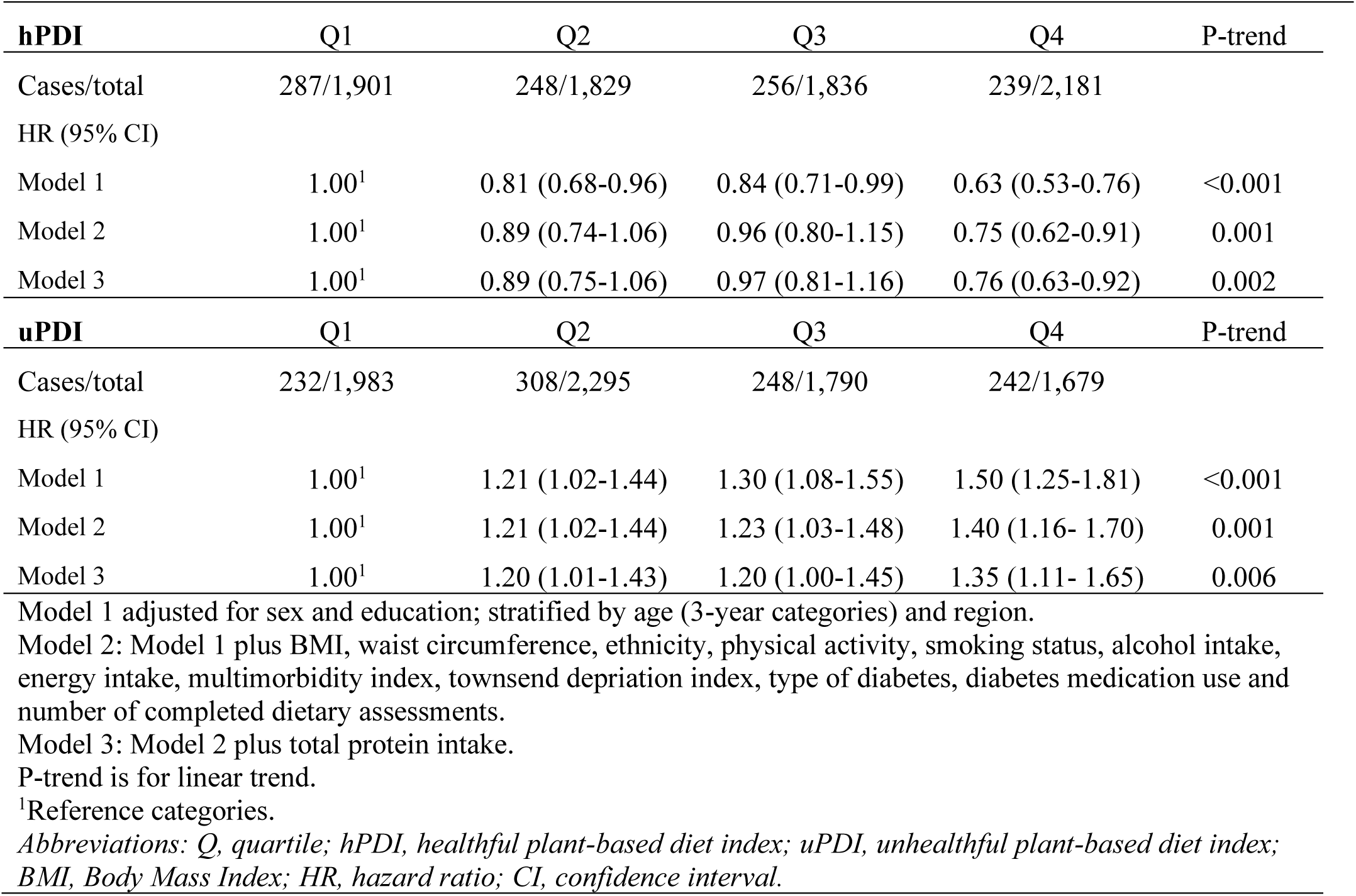
Hazard ratios (95% confidence intervals) of chronic kidney disease among all individuals with diabetes across sex-specific quartiles (Q) of the healthful plant-based diet index (hPDI) and unhealthful plant-based diet index (uPDI) (n=7,747)

| **hPDI** | **Q1** | **Q2** | **Q3** | **Q4** | **P-trend** |
| --- | --- | --- | --- | --- | --- |
| Cases/total | 287/1,901 | 248/1,829 | 256/1,836 | 239/2,181 |  |
| HR (95% CI) |  |  |  |  |  |
| Model 1 | 1.00<sup>1</sup> | 0.81 (0.68-0.96) | 0.84 (0.71-0.99) | 0.63 (0.53-0.76) | <0.001 |
| Model 2 | 1.00<sup>1</sup> | 0.89 (0.74-1.06) | 0.96 (0.80-1.15) | 0.75 (0.62-0.91) | 0.001 |
| Model 3 | 1.00<sup>1</sup> | 0.89 (0.75-1.06) | 0.97 (0.81-1.16) | 0.76 (0.63-0.92) | 0.002 |
| **uPDI** | **Q1** | **Q2** | **Q3** | **Q4** | **P-trend** |
| Cases/total | 232/1,983 | 308/2,295 | 248/1,790 | 242/1,679 |  |
| HR (95% CI) |  |  |  |  |  |
| Model 1 | 1.00<sup>1</sup> | 1.21 (1.02-1.44) | 1.30 (1.08-1.55) | 1.50 (1.25-1.81) | <0.001 |
| Model 2 | 1.00<sup>1</sup> | 1.21 (1.02-1.44) | 1.23 (1.03-1.48) | 1.40 (1.16- 1.70) | 0.001 |
| Model 3 | 1.00<sup>1</sup> | 1.20 (1.01-1.43) | 1.20 (1.00-1.45) | 1.35 (1.11- 1.65) | 0.006 |

Model 1 adjusted for sex and education; stratified by age (3-year categories) and region.
Model 2: Model 1 plus BMI, waist circumference, ethnicity, physical activity, smoking status, alcohol intake, energy intake, multimorbidity index, townsend deprivation index, type of diabetes, diabetes medication use and number of completed dietary assessments.
Model 3: Model 2 plus total protein intake.
P-trend is for linear trend.
<sup>1</sup>Reference categories.
Abbreviations: Q, quartile; hPDI, healthful plant-based diet index; uPDI, unhealthy plant-based diet index; BMI, Body Mass Index; HR, hazard ratio; CI, confidence interval.

In contrast, after multivariable adjustments (model 2), participants with higher uPDI scores (Q4) had a 40% higher risk of CKD (HR: 1.39 [95%CI: 1.16-1.70], p_trend_ = 0.001) compared to those with lower scores (Q1)). Risk estimates were slightly attenuated after further adjusting for total protein intake (model 3), with higher uPDI scores (Q4) being associated with a 35% higher risk of CKD (HR: 1.35 [95%CI: 1.11-1.65], p_trend_ = 0.006) compared to lower scores (Q1).

On the food group level, higher intakes (Q4) of SSBs were associated with a 24% increased risk of CKD (HR: 1.24 [95%CI: 1.05-1.47], p_trend_ = 0.04) compared to lower intakes (Q1). An inverse trend was shown for tea and coffee intake (p_trend_ = 0.02), while a positive trend was evident for meat intake and CKD risk (p_trend_ = 0.02) (**Table 3**).

**Table 3.**
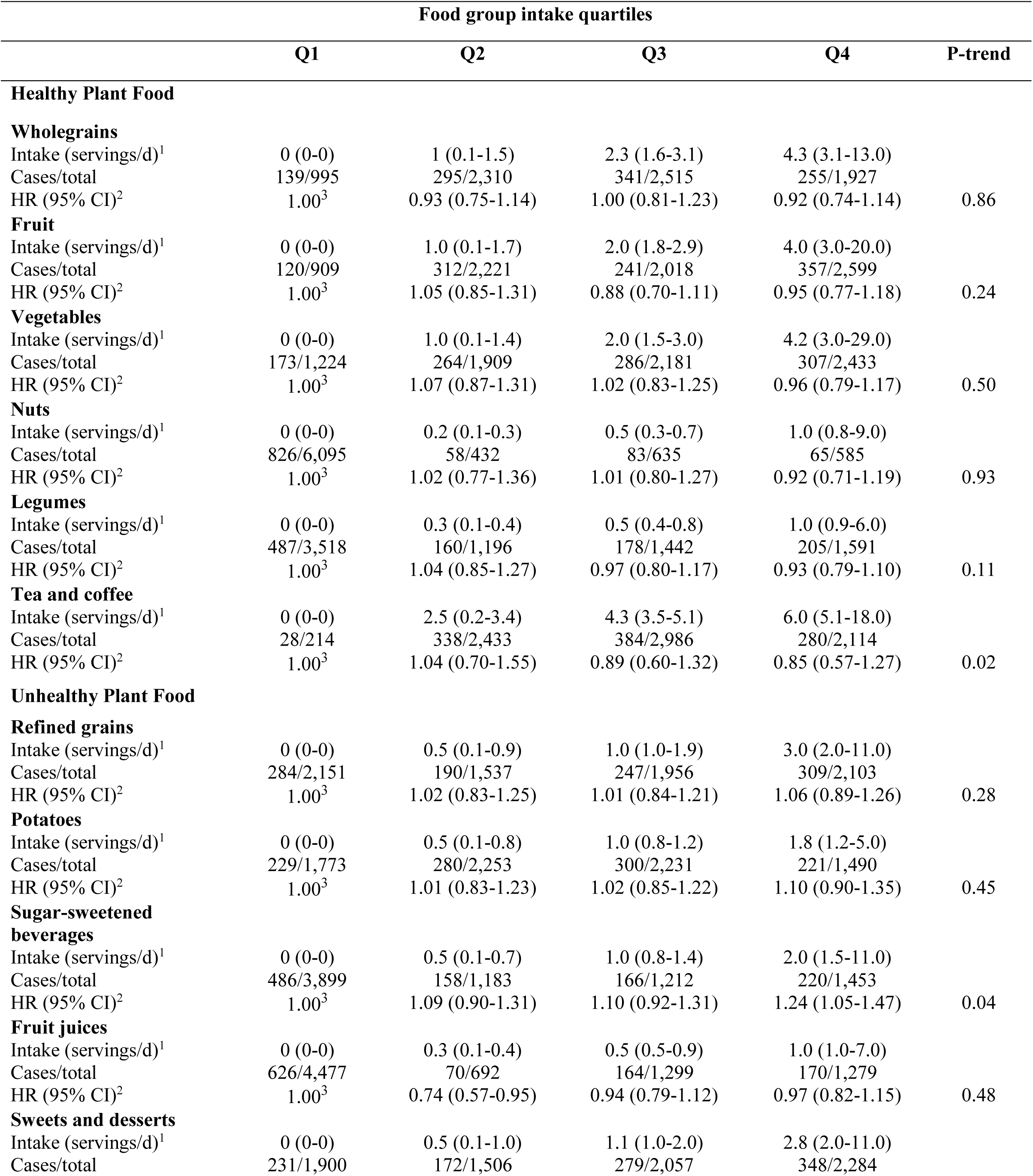
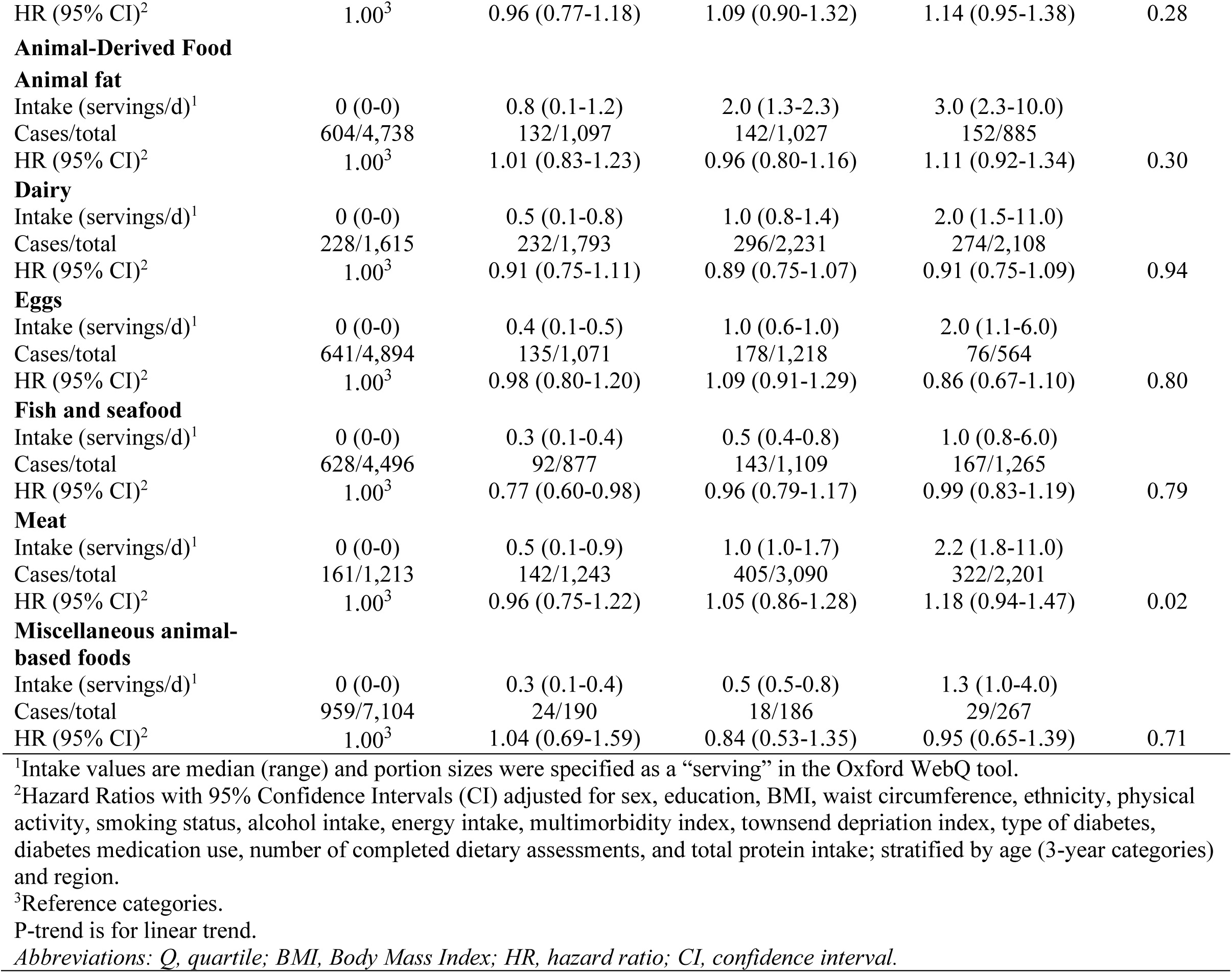
Hazard ratios (95% confidence intervals) of chronic kidney disease across key food groups from the plant-based diet index (n=7,747)

### 3.3 Subgroup and sensitivity analyses

In stratified analyses, there was no indication of heterogeneity across potential effect modifiers (**Supplementary Table S6**). In sensitivity analyses, further adjusting for eGFR, laboratory measurements (LDL-C, HDL-C, lipoprotein(a), C-reactive protein, albumin) and prevalent nephrolithiasis only marginally affected the associations between hPDI (Q4 vs Q1) and CKD risk (**Supplementary Table S7**). Further adjusting for genetic susceptibility of kidney diseases did not notably alter results for either hPDI or uPDI and risk of CKD (**Supplementary Table S8**). Results also remained unchanged on excluding participants who were diagnosed with CKD within the first 2 years of follow-up (**Supplementary Table S9**). Statistically significant associations also persisted on excluding SSBs from the score (**Supplementary Table S10)**.

After employing the E-value methodology in our primary analyses, we found that the E-values were substantially greater than the Hazard ratios observed in fully adjusted models. This suggests that it is unlikely for unmeasured confounding to nullify the significant associations observed. For detailed information, the E-values and upper confidence limits for the fully adjusted main analyses can be found in **Supplementary Tables S12** and **S13.**

### 3.4 Mediation analyses

Among mediation analyses, cystatin C was shown to mediate the observed associations between hPDI and CKD most strongly (proportion mediated: 47%). In addition, creatinine, waist circumference and eGFR showed mediating effects of 39%, 25%, 23% respectively, followed by BMI, triglycerides and urate (6% each), and HDL-C (5%), although p-values for these indirect effects were borderline non-significant (**Table 4**). For uPDI and CKD, cystatin C again showed to mediate the observed associations most strongly (89%), followed by creatinine (44%), waist circumference (25%) and triglycerides (11%). BMI, HDL-C and eGFR also showed to mediate the observed associations between uPDI and CKD by 9-13%, although indirect effects were non-significant (**Supplementary Table S11**). Other potential mediators (including LDL-C, lipoprotein(a), apolipoprotein(a), nephrolithiasis, prevalent hypertension, systolic blood pressure and CRP) were also considered in these analyses but demonstrated no statistically mediating effects.

**Table 4.** Mediation analysis between healthful plant-based diet score and chronic kidney disease.

|  | Healthful Plant-Based Diet Index Score (1-point increments) |  |  |  |  |  |  |  |
| --- | --- | --- | --- | --- | --- | --- | --- | --- |
|  | Participants,<br>No. | Total effect<br>(HR; 95% CI)<sup>1</sup> | *P*-value | Direct effect<br>(HR; 95% CI)<sup>1</sup> | *P*-value | Natural indirect<br>effect<br>(HR; 95% CI)<sup>1</sup> | *P*-value | Proportion mediated<br>(Log(NIE) /<br>(Log(NIE)+log(NDE)<br>) |
| **Potential**<br>**mediators<sup>2</sup>** |  |  |  |  |  |  |  |  |
| **Obesity and lipid**<br>**metabolism** |  |  |  |  |  |  |  |  |
| BMI | 4,756 | 0.982 [0.968-0.997] | 0.022 | 0.984 [0.969-0.999] | 0.037 | 0.999 [0.997-1.000] | 0.070 | 6% |
| Waist<br>circumference | 4,756 | 0.980 [0.965-0.995] | 0.008 | 0.985 [0.970-1.000] | 0.047 | 0.995 [0.993-0.997] | <0.001 | 25% |
| LDL Cholesterol | 4,744 | 0.983 [0.968-0.998] | 0.029 | 0.983 [0.969-0.999] | 0.032 | 1.000 [0.999-1.000] | 0.362 | NA |
| HDL Cholesterol | 4,361 | 0.982 [0.966-0.997] | 0.022 | 0.982 [0.967-0.998] | 0.030 | 0.999 [0.998-1.000] | 0.066 | 5% |
| Triglycerides | 4,753 | 0.983 [0.968-0.998] | 0.030 | 0.984 [0.969-0.999] | 0.037 | 0.999 [0.999-1.000] | 0.079 | 6% |
| Lipoprotein A | 3,639 | 0.979 [0.962-0.996] | 0.015 | 0.979 [0.962-0.996] | 0.016 | 1.000 [0.999-1.000] | 0.579 | NA |
| Apolipoprotein A | 4,351 | 0.981 [0.966-0.997] | 0.021 | 0.982 [0.966-0.997] | 0.023 | 1.000 [0.999-1.000] | 0.334 | NA |
| **Kidney function** |  |  |  |  |  |  |  |  |
| eGFR | 4,756 | 0.983 [0.967-0.999] | 0.034 | 0.987 [0.972-1.003] | 0.105 | 0.996 [0.993-0.999] | 0.007 | 23% |
| Cystatin C | 4,754 | 0.982 [0.967-0.998] | 0.032 | 0.991 [0.975-1.007] | 0.255 | 0.992 [0.988-0.995] | <0.001 | 47% |
| Urate | 4,748 | 0.982 [0.967-0.998] | 0.025 | 0.984 [0.969-0.999] | 0.038 | 0.999 [0.997-1.000] | 0.106 | 6% |
| Creatinine | 4,756 | 0.982 [0.967-0.998] | 0.027 | 0.989 [0.973-1.004] | 0.154 | 0.993 [0.990-0.997] | <0.001 | 39% |
| Nephrolithiasis | 4,756 | 0.983 [0.968-0.998] | 0.030 | 0.983 [0.968-0.998] | 0.029 | 1.000 [0.999-1.000] | 0.962 | NA |
| **Hypertension** |  |  |  |  |  |  |  |  |
| Prevalent<br>hypertension | 4,756 | 0.983 [0.967-1.000] | 0.045 | 0.983 [0.969-0.998] | 0.030 | 1.000 [0.994-1.006] | 0.999 | NA |
| Systolic blood<br>pressure | 4,601 | 0.982 [0.967-0.997] | 0.019 | 0.982 [0.967-0.997] | 0.018 | 1.000 [0.999-1.001] | 0.655 | NA |

| Inflammatory biomarkers |  |  |  |  |  |  |  |  |
| --- | --- | --- | --- | --- | --- | --- | --- | --- |
| C-reactive protein | 4,746 | 0.983 [0.968-0.998] | 0.029 | 0.984 [0.969-0.999] | 0.033 | 1.000 [0.999-1.000] | 0.551 | NA |
| <sup>1</sup>Hazard Ratios with 95% Confidence Intervals (CI) for healthful plant-based diet score (1-point increments), adjusted for sex, BMI (excluding when considered as a potential mediator), waist circumference (excluding when considered as a potential mediator), ethnicity, physical activity, smoking status, education, alcohol intake, energy intake, multimorbidity index, Townsend deprivation index, type of diabetes, diabetes medication use, number of completed dietary assessments, and total protein intake; stratified by age (3-year categories) and region.<br><sup>2</sup>Potential mediators are modelled on the continuous scale (excluding prevalent nephrolithiasis and prevalent hypertension which are binary).<br>Abbreviations: *HR*, hazard ratios; *CI*, confidence intervals; *BMI*, body mass index; *eGFR*, estimated glomerular filtration rate; *NIE*, natural indirect effect; *NDE*, natural direct effect. |  |  |  |  |  |  |  |  |

## 4. Discussion

In this cohort of UK adults living with diabetes, greater adherence to a healthy PBD was associated with a 24% lower risk of CKD, while greater adherence to an unhealthy PBD was associated with a 35% higher risk. The associations between a healthy PBD and CKD were primarily mediated by improved kidney function (specifically by cystatin C) and lower body fatness.

To the best of our knowledge, this is the first prospective study to examine the associations between a healthy and unhealthy PBD and risk of CKD among middle-aged adults living with diabetes. Our results are in line with a retrospective study of 2,797 diabetic patients from Taiwan where they showed that vegetarian participants had a lower risk of CKD compared to omnivores (39), and a cross-sectional analysis in 420 diabetic patients from the Netherlands which showed higher vegetable protein intake to be associated with improved renal function compared to lower intakes (40). While our findings add to the growing body of evidence that dietary patterns rich in plant foods improve outcomes among people with diabetes (5), they also align with those from meta-analyses on healthy dietary patterns and CKD incidence in people with and without diabetes. These studies show a diet rich in whole grains, vegetables, fruit, legumes, nuts, and fish, and a lower intake of red and processed meats, sodium, and SSBs to be associated with lower CKD risk and albuminuria (41, 42).

In our study, the overall quality of PBDs was associated with CKD risk, while analyses on the food group level showed SSBs to be most strongly associated with increased CKD risk, which is in line with previous studies (43). Several plausible mechanisms may explain the associations found between SSBs and CKD in our study. Firstly, SSBs are typically energy-dense and, when overconsumed, can contribute to weight gain, overweight and obesity (44) – a known risk factor for new-onset kidney disease (45). Further, SSBs have a high glycaemic index (46) which may increase glucose and insulin levels, both of which can impair renal function through chronic low-grade inflammation and hyperglycaemia (47, 48). Additionally, fructose, which is a main component of SSBs, is associated with hyperuricemia and increased renin expression, which in turn can accelerate the onset of renal disease and progression through vascular disease and interstitial fibrosis (49, 50). Fructose has also been independently associated with kidney stone incidence (51), which is an established CKD risk factor associated with adverse renal outcomes (37, 52). Of note, however, the associations in our study remained similar upon excluding SSBs from the PBD scores, which speaks for other benefits of healthy PBDs beyond lower SSBs consumption.

The beneficial effects of a healthy PBD on kidney function may also be explained through greater dietary fibre intake. Although no associations were found between whole grain intake and CKD risk in our study, we found that those in the highest quartile of hPDI had greater overall dietary fibre intake. In the Tehran Lipid and Glucose study (n=1,630), a 5 g/day increase in total fibre was associated with an 11% lower risk of CKD (53), while higher dietary fibre intake was associated with a 24% lower risk of incident CKD in a recent study of the UK Biobank cohort (54). Mechanisms explaining the beneficial associations of dietary fibre on CKD risk include its cholesterol-lowering (55, 56) and blood pressure lowering (57) attributes. Further, dietary fibre improves glycaemic control and insulin secretion, which significantly lowers the risk of developing microalbuminuria and proteinuria (58). In our study of participants with diabetes, improved glycaemic control through higher dietary fibre intake may in part explain the benefit of a healthy PBD on CKD risk. Dietary fibre intake can delay gastric emptying, slow post-meal glucose absorption, and increase satiety improving overall insulin sensitivity, which may in turn delay the onset and progression of CKD (59–61).

While our goal was to investigate PBDs in relation to CKD risk among people with diabetes, there is one previous similar study conducted in the general population (9). Specifically, data from a U.S cohort of healthy adults found that higher adherence to a healthy plant-based dietary pattern was associated with a 14% lower risk of CKD and slower eGFR decline, while higher adherence to a less healthy plant-based dietary pattern was associated with an 11% higher risk of CKD. Unlike this study, our analyses considered total protein intake as a potential confounder because dietary protein intake may affect kidney health (62–65). Further, those following an animal-based diet typically have a higher protein intake, making total protein intake an important confounding factor to consider (66). A prospective study recently carried out on plant protein intake and incident CKD demonstrated that those in the highest quartile of intake had an 18% lower risk of CKD compared with the lowest quartile of intake (10). Our findings suggest that a healthy PBD is beneficial for lowering CKD risk irrespective of protein intake. This is potentially because healthy PBDs are typically of higher nutritional value, providing a rich source of micronutrients which have been shown to improve kidney function through lowering oxidative stress, inflammation, dietary acid load and endothelial dysfunction (67, 68). There are no randomized controlled trials on plant protein intake and incident CKD to support these observational findings, however, results from 3 small human trials in patients with albuminuria and type 2 diabetes found a statistically significant reduction in albuminuria upon replacing animal protein with plant protein, controlling for total protein intake (69–71).

### 4.1 Limitations

Although this study has many strengths including its large sample size, prospective design, and substantial follow-up time, there are also some limitations to consider. Firstly, while the observed associations were obtained after meticulous adjustment for potential confounding factors, we cannot rule out residual confounding. Secondly, to utilise the maximum number of patients with diabetes and 24-hr dietary data in this cohort, the PDI was estimated using dietary data from participants who completed a minimum of one 24-h dietary assessment. Therefore, it is possible that the PDI does not fully represent participants’ habitual dietary intake (72). Despite this, the PDIs have previously been shown to have good reproducibility over time (73). Thirdly, hospital inpatient and death registry data were used to ascertain incident CKD cases, but not primary care data. Linkage to primary care data in the UK Biobank is currently ongoing (data is available for approximately 45% of UK Biobank cohort) and was therefore not used for this analysis. Finally, the UK Biobank cohort is made up of British middle-aged adults, predominantly of which are of White European ancestry (>90%), higher education level, and live a healthier lifestyle (74); all of which may limit the generalizability of our findings to other non-European and ethnic populations.

## 5. Conclusion

Findings from this cohort study of 7,747 middle-aged adults with diabetes suggest that a healthy PBD is associated with a 24% lower CKD risk irrespective of established CKD risk factors, genetic susceptibility to kidney disease and total protein intake. In contrast, an unhealthy PBD was associated with a 35% higher risk of CKD. Markers of kidney function, obesity, lipid metabolism and inflammation were found to potentially mediate the observed associations between plant-based dietary intake and CKD. These findings highlight the potential importance of PBD quality in the form of higher intakes of healthy plant foods and lower intakes of unhealthy plant foods to lower CKD risk among those living with diabetes. In addition, our findings suggest that SSBs should be restricted for the prevention of CKD in diabetic patients. RCTs would be an important next step to confirm the role of PBDs in CKD prevention among people with diabetes.

## Data Availability

UK Biobank data can be requested by all bona fide researchers for approved projects, including replication, through https://www.ukbiobank.ac.uk/. This research was conducted using UK Biobank funded and sourced data (application 64426).

## Authors’ contributions

Design and concept: AT, TK, AC; database development: AT, ATR, JKO, CH; analysed and interpreted data: AT, AC, TK; drafted manuscript: AT, TK, AC; provided critical review of the manuscript: ATR, AJ, NB, CC, JO, CH, MG; guarantors of the work: AT and TK.

## Funding and Acknowledgements

This research was conducted using UK Biobank funded and sourced data (application 64426). The UK Biobank was established by the Wellcome Trust, the Medical Research Council, the UK Department of Health, and the Scottish Government. The UK Biobank has also received funding from the Welsh Assembly Government, the British Heart Foundation, and Diabetes United Kingdom, Northwest Regional Development Agency, Scottish Government. In addition, Alysha S. Thompson holds a PhD studentship of the Department for the Economy (DfE), Northern Ireland.

## Ethics approval

All UK Biobank participants provided informed consent to participate and be followed through linkage to their health records. The UK Biobank study received ethical approval from the NHS North West Multicentre Research Ethics Committee (Ref. 11/NW/0382).

## References

1. Bikbov B, Purcell CA, Levey AS, Smith M, Abdoli A, Abebe M, et al. Global, regional, and national burden of chronic kidney disease, 1990–2017: a systematic analysis for the Global Burden of Disease Study 2017. The lancet. 2020;395(10225):709–33.

2. Couser WG, Remuzzi G, Mendis S, Tonelli M. The contribution of chronic kidney disease to the global burden of major noncommunicable diseases. Kidney international. 2011;80(12):1258–70.

3. Levin A, Tonelli M, Bonventre J, Coresh J, Donner J-A, Fogo AB, et al. Global kidney health 2017 and beyond: a roadmap for closing gaps in care, research, and policy. The Lancet. 2017;390(10105):1888–917.

4. Eckardt K-U, Coresh J, Devuyst O, Johnson RJ, Köttgen A, Levey AS, et al. Evolving importance of kidney disease: from subspecialty to global health burden. The Lancet. 2013;382(9887):158–69.

5. Whiteley C, Benton F, Matwiejczyk L, Luscombe-Marsh N. Determining Dietary Patterns to Recommend for Type 2 Diabetes: An Umbrella Review. Nutrients. 2023;15(4):861. PubMed PMID: doi:10.3390/nu15040861.

6. Xu S-S, Hua J, Huang Y-Q, Shu L. Association between dietary patterns and chronic kidney disease in a middle-aged Chinese population. Public Health Nutr. 2020;23(6):1058–66.

7. Gu Y, Li H, Ma H, Zhang S, Meng G, Zhang Q, et al. Consumption of ultraprocessed food and development of chronic kidney disease: the Tianjin Chronic Low-Grade Systemic Inflammation and Health and UK Biobank Cohort Studies. The American Journal of Clinical Nutrition. 2023;117(2):373–82. doi: 10.1016/j.ajcnut.2022.11.005.

8. Du S, Kim H, Crews DC, White K, Rebholz CM. Association Between Ultraprocessed Food Consumption and Risk of Incident CKD: A Prospective Cohort Study. American Journal of Kidney Diseases. 2022;80(5):589–98.e1. doi: 10.1053/j.ajkd.2022.03.016.

9. Kim H, Caulfield LE, Garcia-Larsen V, Steffen LM, Grams ME, Coresh J, et al. Plant-Based Diets and Incident CKD and Kidney Function. Clin J Am Soc Nephrol. 2019;14(5):682–91. Epub 2019/04/27. doi: 10.2215/cjn.12391018. PubMed PMID: 31023928; PubMed Central PMCID: PMCPMC6500948.

10. Heo GY, Koh HB, Kim HJ, Kim KW, Jung C-Y, Kim HW, et al. Association of Plant Protein Intake With Risk of Incident CKD: A UK Biobank Study. American Journal of Kidney Diseases. 2023. doi: 10.1053/j.ajkd.2023.05.007.

11. Haring B, Selvin E, Liang M, Coresh J, Grams ME, Petruski-Ivleva N, et al. Dietary protein sources and risk for incident chronic kidney disease: results from the Atherosclerosis Risk in Communities (ARIC) Study. Journal of Renal Nutrition. 2017;27(4):233–42.

12. Rossing P, Caramori ML, Chan JC, Heerspink HJ, Hurst C, Khunti K, et al. KDIGO 2022 clinical practice guideline for diabetes management in chronic kidney disease. Kidney International. 2022;102(5):S1–S127.

13. Sudlow C, Gallacher J, Allen N, Beral V, Burton P, Danesh J, et al. UK biobank: an open access resource for identifying the causes of a wide range of complex diseases of middle and old age. PLoS medicine. 2015;12(3):e1001779–e. doi: 10.1371/journal.pmed.1001779. PubMed PMID: 25826379.

14. Von Elm E, Altman DG, Egger M, Pocock SJ, Gøtzsche PC, Vandenbroucke JP. The Strengthening the Reporting of Observational Studies in Epidemiology (STROBE) statement: guidelines for reporting observational studies. The Lancet. 2007;370(9596):1453–7.

15. Schofield WN, Schofield C, James WPT. Basal metabolic rate: review and prediction, together with an annotated bibliography of source material. 1985.

16. Perez-Cornago A, Pollard Z, Young H, van Uden M, Andrews C, Piernas C, et al. Description of the updated nutrition calculation of the Oxford WebQ questionnaire and comparison with the previous version among 207,144 participants in UK Biobank. European Journal of Nutrition. 2021;60(7):4019–30. doi: 10.1007/s00394-021-02558-4.

17. Greenwood DC, Hardie LJ, Frost GS, Alwan NA, Bradbury KE, Carter M, et al. Validation of the Oxford WebQ Online 24-Hour Dietary Questionnaire Using Biomarkers. Am J Epidemiol. 2019;188(10):1858–67. doi: 10.1093/aje/kwz165.

18. Galante J, Adamska L, Young A, Young H, Littlejohns TJ, Gallacher J, et al. The acceptability of repeat Internet-based hybrid diet assessment of previous 24-h dietary intake: administration of the Oxford WebQ in UK Biobank. British Journal of Nutrition. 2016;115(4):681–6.

19. Liu B, Young H, Crowe FL, Benson VS, Spencer EA, Key TJ, et al. Development and evaluation of the Oxford WebQ, a low-cost, web-based method for assessment of previous 24 h dietary intakes in large-scale prospective studies. Public Health Nutr. 2011;14(11):1998–2005. Epub 2011/06/01. doi: 10.1017/S1368980011000942.

20. Satija A, Bhupathiraju SN, Rimm EB, Spiegelman D, Chiuve SE, Borgi L, et al. Plant-Based Dietary Patterns and Incidence of Type 2 Diabetes in US Men and Women: Results from Three Prospective Cohort Studies. PLOS Medicine. 2016;13(6):e1002039. doi: 10.1371/journal.pmed.1002039.

21. Satija A, Bhupathiraju SN, Spiegelman D, Chiuve SE, Manson JE, Willett W, et al. Healthful and Unhealthful Plant-Based Diets and the Risk of Coronary Heart Disease in U.S. Adults. Journal of the American College of Cardiology. 2017;70(4):411–22. doi: 10.1016/j.jacc.2017.05.047.

22. Heianza Y, Zhou T, Sun D, Hu FB, Qi L. Healthful plant-based dietary patterns, genetic risk of obesity, and cardiovascular risk in the UK biobank study. Clinical Nutrition. 2021;40(7):4694–701.

23. Piernas C, Perez-Cornago A, Gao M, Young H, Pollard Z, Mulligan A, et al. Describing a new food group classification system for UK biobank: analysis of food groups and sources of macro-and micronutrients in 208,200 participants. European Journal of Nutrition. 2021;60:2879–90.

24. Eastwood SV, Mathur R, Atkinson M, Brophy S, Sudlow C, Flaig R, et al. Algorithms for the capture and adjudication of prevalent and incident diabetes in UK Biobank. PloS one. 2016;11(9):e0162388.

25. Levey AS, Stevens LA, Schmid CH, Zhang Y, Castro III AF, Feldman HI, et al. A new equation to estimate glomerular filtration rate. Annals of internal medicine. 2009;150(9):604–12.

26. Wuttke M, Li Y, Li M, Sieber KB, Feitosa MF, Gorski M, et al. A catalog of genetic loci associated with kidney function from analyses of a million individuals. Nature genetics. 2019;51(6):957–72.

27. VanderWeele TJ, Ding P. Sensitivity analysis in observational research: introducing the E-value. Annals of internal medicine. 2017;167(4):268–74.

28. Emsley RA, Liu H, Dunn G, Valeri L, VanderWeele TJ. Paramed: A command to perform causal mediation analysis using parametric models. The Stata Journal. 2014.

29. Dunn G, Emsley R, Liu H, Landau S, Green J, White I, et al. Evaluation and validation of social and psychological markers in randomised trials of complex interventions in mental health: a methodological research programme. Health Technology Assessment (Winchester, England). 2015;19(93):1–116.

30. Fox CS, Larson MG, Leip EP, Culleton B, Wilson PW, Levy D. Predictors of new-onset kidney disease in a community-based population. Jama. 2004;291(7):844–50.

31. Liu L, Wang Y, Zhang W, Chang W, Jin Y, Yao Y. Waist height ratio predicts chronic kidney disease: a systematic review and meta-analysis, 1998-2019. Arch Public Health. 2019;77:55. Epub 2019/12/24. doi: 10.1186/s13690-019-0379-4. PubMed PMID: 31867106; PubMed Central PMCID: PMCPMC6918668.

32. Bae JC, Han JM, Kwon S, Jee JH, Yu TY, Lee MK, et al. LDL-C/apoB and HDL-C/apoA-1 ratios predict incident chronic kidney disease in a large apparently healthy cohort. Atherosclerosis. 2016;251:170–6. doi: 10.1016/j.atherosclerosis.2016.06.029.

33. Eknoyan G, Lameire N, Eckardt K, Kasiske B, Wheeler D, Levin A, et al. KDIGO 2012 clinical practice guideline for the evaluation and management of chronic kidney disease. Kidney int. 2013;3(1):5–14.

34. Benoit SW, Ciccia EA, Devarajan P. Cystatin C as a biomarker of chronic kidney disease: latest developments. Expert review of molecular diagnostics. 2020;20(10):1019–26.

35. Kang D-H, Nakagawa T, Feng L, Watanabe S, Han L, Mazzali M, et al. A role for uric acid in the progression of renal disease. Journal of the American Society of Nephrology. 2002;13(12):2888–97.

36. Zhang WR, Parikh CR. Biomarkers of acute and chronic kidney disease. Annual review of physiology. 2019;81:309–33.

37. Rule AD, Bergstralh EJ, Melton III LJ, Li X, Weaver AL, Lieske JC. Kidney stones and the risk for chronic kidney disease. Clinical journal of the American Society of Nephrology: CJASN. 2009;4(4):804.

38. Kugler E, Cohen E, Goldberg E, Nardi Y, Levi A, Krause I, et al. C reactive protein and long-term risk for chronic kidney disease: a historical prospective study. Journal of Nephrology. 2015;28(3):321–7. doi: 10.1007/s40620-014-0116-6.

39. Hou YC, Huang HF, Tsai WH, Huang SY, Liu HW, Liu JS, et al. Vegetarian Diet Was Associated With a Lower Risk of Chronic Kidney Disease in Diabetic Patients. Front Nutr. 2022;9:843357. Epub 2022/05/14. doi: 10.3389/fnut.2022.843357. PubMed PMID: 35558755; PubMed Central PMCID: PMCPMC9087577.

40. Oosterwijk MM, Soedamah-Muthu SS, Geleijnse JM, Bakker SJ, Navis G, Binnenmars SH, et al. High dietary intake of vegetable protein is associated with lower prevalence of renal function impairment: results of the Dutch DIALECT-1 cohort. Kidney international reports. 2019;4(5):710–9.

41. Bach KE, Kelly JT, Palmer SC, Khalesi S, Strippoli GFM, Campbell KL. Healthy Dietary Patterns and Incidence of CKD: A Meta-Analysis of Cohort Studies. Clin J Am Soc Nephrol. 2019;14(10):1441–9. Epub 2019/09/26. doi: 10.2215/cjn.00530119. PubMed PMID: 31551237; PubMed Central PMCID: PMCPMC6777603.

42. He L-Q, Wu X-H, Huang Y-Q, Zhang X-Y, Shu L. Dietary patterns and chronic kidney disease risk: a systematic review and updated meta-analysis of observational studies. Nutrition Journal. 2021;20(1):4. doi: 10.1186/s12937-020-00661-6.

43. Lo W-C, Ou S-H, Chou C-L, Chen J-S, Wu M-Y, Wu M-S. Sugar- and artificially-sweetened beverages and the risks of chronic kidney disease: a systematic review and dose– response meta-analysis. Journal of Nephrology. 2021;34(6):1791–804. doi: 10.1007/s40620-020-00957-0.

44. Malik VS, Pan A, Willett WC, Hu FB. Sugar-sweetened beverages and weight gain in children and adults: a systematic review and meta-analysis. The American journal of clinical nutrition. 2013;98(4):1084–102.

45. Kovesdy CP, Furth S, Zoccali C, Committee WKDS. Obesity and kidney disease: hidden consequences of the epidemic. Physiology international. 2017;104(1):1–14.

46. Atkinson FS, Foster-Powell K, Brand-Miller JC. International tables of glycemic index and glycemic load values: 2008. Diabetes care. 2008;31(12):2281–3.

47. Gopinath B, Harris DC, Flood VM, Burlutsky G, Brand-Miller J, Mitchell P. Carbohydrate Nutrition Is Associated with the 5-Year Incidence of Chronic Kidney Disease. The Journal of Nutrition. 2011;141(3):433–9. doi: 10.3945/jn.110.134304.

48. Liu S, Manson JE, Buring JE, Stampfer MJ, Willett WC, Ridker PM. Relation between a diet with a high glycemic load and plasma concentrations of high-sensitivity C-reactive protein in middle-aged women. The American journal of clinical nutrition. 2002;75(3):492–8.

49. Zheng Z, Harman JL, Coresh J, Köttgen A, McAdams-DeMarco MA, Correa A, et al. The dietary fructose: vitamin C intake ratio is associated with hyperuricemia in african-american adults. The Journal of Nutrition. 2018;148(3):419–26.

50. Johnson RJ, Segal MS, Sautin Y, Nakagawa T, Feig DI, Kang D-H, et al. Potential role of sugar (fructose) in the epidemic of hypertension, obesity and the metabolic syndrome, diabetes, kidney disease, and cardiovascular disease. The American journal of clinical nutrition. 2007;86(4):899–906.

51. Asselman M, Verkoelen C. Fructose intake as a risk factor for kidney stone disease. Kidney International. 2008;73(2):139–40.

52. Alexander RT, Hemmelgarn BR, Wiebe N, Bello A, Morgan C, Samuel S, et al. Kidney stones and kidney function loss: a cohort study. Bmj. 2012;345.

53. Mirmiran P, Yuzbashian E, Asghari G, Sarverzadeh S, Azizi F. Dietary fibre intake in relation to the risk of incident chronic kidney disease. British Journal of Nutrition. 2018;119(5):479–85.

54. Heo GY, Kim HJ, Kalantar D, Jung CY, Kim HW, Park JT, et al. Association between Fiber Intake and Risk of Incident Chronic Kidney Disease: The UK Biobank Study. The journal of nutrition, health & aging. 2023;27(11):1018–27. doi: 10.1007/s12603-023-1998-6.

55. Chutkan R, Fahey G, Wright WL, McRorie J. Viscous versus nonviscous soluble fiber supplements: Mechanisms and evidence for fiber-specific health benefits. Journal of the American Academy of Nurse Practitioners. 2012;24(8):476–87.

56. Trautwein EA, McKay S. The Role of Specific Components of a Plant-Based Diet in Management of Dyslipidemia and the Impact on Cardiovascular Risk. Nutrients. 2020;12(9). Epub 2020/09/05. doi: 10.3390/nu12092671. PubMed PMID: 32883047; PubMed Central PMCID: PMCPMC7551487.

57. Streppel MT, Arends LR, van’t Veer P, Grobbee DE, Geleijnse JM. Dietary fiber and blood pressure: a meta-analysis of randomized placebo-controlled trials. Archives of internal medicine. 2005;165(2):150–6.

58. Fioretto P, Bruseghin M, Berto I, Gallina P, Manzato E, Mussap M. Renal protection in diabetes: role of glycemic control. Journal of the American Society of Nephrology. 2006;17(4_suppl_2):S86–S9.

59. Shurraw S, Hemmelgarn B, Lin M, Majumdar SR, Klarenbach S, Manns B, et al. Association between glycemic control and adverse outcomes in people with diabetes mellitus and chronic kidney disease: a population-based cohort study. Archives of internal medicine. 2011;171(21):1920–7.

60. Kalantar-Zadeh K, Rhee CM, Joshi S, Brown-Tortorici A, Kramer HM. Medical nutrition therapy using plant-focused low-protein meal plans for management of chronic kidney disease in diabetes. Current opinion in nephrology and hypertension. 2022;31(1):26–35.

61. Tuttle KR, Bakris GL, Bilous RW, Chiang JL, De Boer IH, Goldstein-Fuchs J, et al. Diabetic kidney disease: a report from an ADA Consensus Conference. Diabetes care. 2014;37(10):2864–83.

62. Knight EL, Stampfer MJ, Hankinson SE, Spiegelman D, Curhan GC. The impact of protein intake on renal function decline in women with normal renal function or mild renal insufficiency. Annals of internal medicine. 2003;138(6):460–7.

63. Lew Q-LJ, Jafar TH, Koh HWL, Jin A, Chow KY, Yuan J-M, et al. Red meat intake and risk of ESRD. Journal of the American Society of Nephrology: JASN. 2017;28(1):304.

64. Lin J, Fung TT, Hu FB, Curhan GC. Association of dietary patterns with albuminuria and kidney function decline in older white women: a subgroup analysis from the Nurses’ Health Study. Am J Kidney Dis. 2011;57(2):245–54. Epub 2011/01/22. doi: 10.1053/j.ajkd.2010.09.027. PubMed PMID: 21251540; PubMed Central PMCID: PMCPMC3026604.

65. Rebholz CM, Crews DC, Grams ME, Steffen LM, Levey AS, Miller III ER, et al. DASH (Dietary Approaches to Stop Hypertension) diet and risk of subsequent kidney disease. American Journal of Kidney Diseases. 2016;68(6):853–61.

66. Clarys P, Deliens T, Huybrechts I, Deriemaeker P, Vanaelst B, De Keyzer W, et al. Comparison of nutritional quality of the vegan, vegetarian, semi-vegetarian, pesco-vegetarian and omnivorous diet. Nutrients. 2014;6(3):1318–32.

67. Mirmiran P, Yuzbashian E, Bahadoran Z, Asghari G, Azizi F. Dietary acid-base load and risk of chronic kidney disease in adults. 2016.

68. Farhadnejad H, Asghari G, Mirmiran P, Yuzbashian E, Azizi F. Micronutrient intakes and incidence of chronic kidney disease in adults: Tehran Lipid and Glucose Study. Nutrients. 2016;8(4):217.

69. Azadbakht L, Esmaillzadeh A. Soy-protein consumption and kidney-related biomarkers among type 2 diabetics: a crossover, randomized clinical trial. Journal of Renal Nutrition. 2009;19(6):479–86.

70. Teixeira SR, Tappenden KA, Carson L, Jones R, Prabhudesai M, Marshall WP, et al. Isolated soy protein consumption reduces urinary albumin excretion and improves the serum lipid profile in men with type 2 diabetes mellitus and nephropathy. The Journal of nutrition. 2004;134(8):1874–80.

71. Azadbakht L, Atabak S, Esmaillzadeh A. Soy protein intake, cardiorenal indices, and C-reactive protein in type 2 diabetes with nephropathy: a longitudinal randomized clinical trial. Diabetes care. 2008;31(4):648–54.

72. Jackson KA, Byrne NM, Magarey AM, Hills AP. Minimizing random error in dietary intakes assessed by 24-h recall, in overweight and obese adults. European journal of clinical nutrition. 2008;62(4):537–43.

73. Thompson AS, Tresserra-Rimbau A, Karavasiloglou N, Jennings A, Cantwell M, Hill C, et al. Association of Healthful Plant-based Diet Adherence With Risk of Mortality and Major Chronic Diseases Among Adults in the UK. JAMA Network Open. 2023;6(3):e234714-e. doi: 10.1001/jamanetworkopen.2023.4714.

74. Fry A, Littlejohns TJ, Sudlow C, Doherty N, Adamska L, Sprosen T, et al. Comparison of sociodemographic and health-related characteristics of UK Biobank participants with those of the general population. Am J Epidemiol. 2017;186(9):1026–34.

