## Supplementary file 1 for "Adherence to a healthful plant-based diet and risk of chronic kidney disease among individuals with diabetes: A prospective cohort study"

**Supplementary file 1 – Adherence to a healthful plant-based diet and risk of chronic kidney disease among individuals with diabetes: A prospective cohort study**

***Thompson et al.***

**Table S7.** Sensitivity analyses showing hazard ratios (95% confidence intervals) across sex-specific healthful vs unhealthful plant-based diet index quartiles (Q), further adjusting for eGFR, laboratory measurements (LDL-C, HDL-C), lipoprotein(a), C-reactive protein, albumin) and nephrolithiasis, for participants who completed 1 or more dietary assessments and the associated risk of chronic kidney disease (n=5,405) ........................................................................................................ 16

**Methods S1:** Genotyping and quality control

Additional quality control (QC) procedures were implemented using PLINK v2.0 (https://www.cog-genomics.org/plink/2.0/) (1, 2). Quality control measures included applying a number of thresholds. 1) All analyses were restricted to individuals with self-reported White British ancestry, with matching self-reported and genetic sex. 2) Analyses were restricted to autosomal variants; excluding samples/variants based on a genotype call rate of ≤95%, an imputation rate of ≤97%, a minor allele frequency of ≤0.001, minor allele minimum count of 5, individual missingness ≤95% and a Hardy-Weinberg equilibrium of ≤1e-8. 3) Outliers for heterozygosity and missingness (PCA corrected), individuals with putative sex aneuploidy and duplicates were also removed using previously defined metrics provided by the UKB (3). The final genetic UKB sample therefore comprised of 408,110 individuals (220,618 females and 187,492 males).

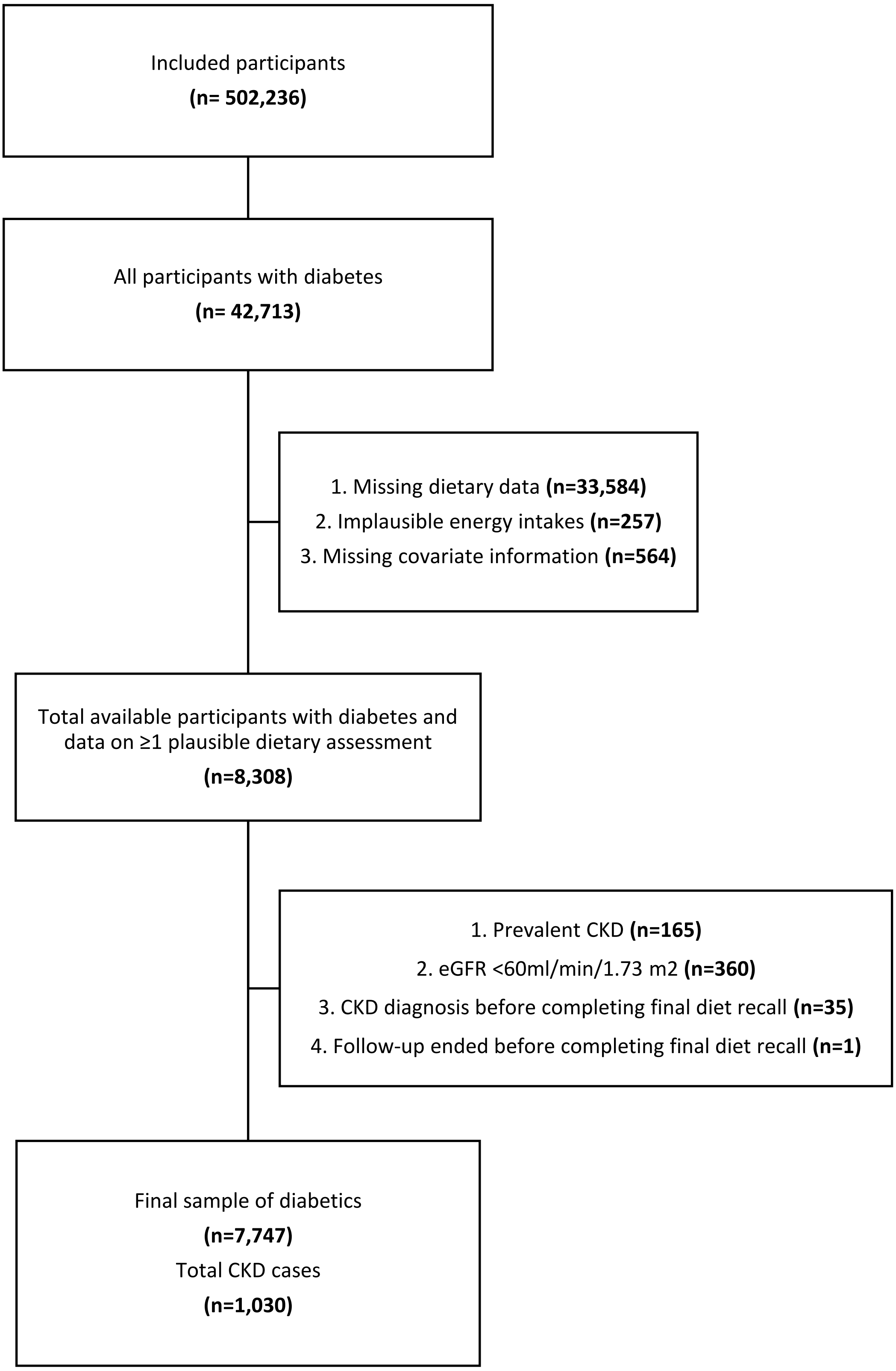

**Figure S1**. Flowchart of exclusions

**Table S1.** Covariate coding and categorisation information

| Variables | Categorisation | UK Biobank variable description and data-field ID |
|---|---|---|
| **Outcome** | | |
| Chronic kidney disease | No; Yes | ICD-10 codes: N03, N06, N08, N11, N12, N13, N14, N15, N16, N18, N19, Z49, I12, I13<br>OPCS-4 codes: L74.1, L74.2, L74.3, L74.4, L74.5, L74.6, L74.8, L74.9, M01.2, M01.3, M01.4, M01.5, M01.8, M01.9, M02.3, M08.4, M17.2, M17.4, M17.8, M17.9, X40.1, X40.2, X40.3, X40.4, X40.5, X40.6, X40.7, X40.8, X40.9, X41.1, X41.2, X41.8, X41.9, X42.1, X42.8, X42.9, and X43.1 |
| **Demographics** | | |
| Age | 3-year categories (<55, 55-<65, ≥65 years) | Age at recruitment (ID: 21022)[1] |
| Sex | Female; Male | Sex (ID: 31)[1] |
| Ethnicity | Asian, Black, Multiple, White, Other/Unknown/Missing | Ethnic background (ID: 21000)[1] |
| Region | London; Wales; North-West England; North-East England; Yorkshire; West Midlands; East Midlands; South-East England; South-West England; Scotland | UK Biobank assessment centre (ID:54)[1] |
| **Socioeconomic status** | | |
| Education | Low: CSEs or equivalent, O levels/GCSEs or equivalent; Medium: A levels/AS levels or equivalent, NVQ or HND or HNC or equivalent; High: College or University degree, other professional qualifications eg: nursing, teaching; Unknown/Missing | Qualifications (ID: 6138)[1] |
| Townsend deprivation index | Quintiles from least to most deprived | Townsend deprivation index (ID: 189)[1] |
| **Diet and Lifestyle** | | |
| Alcohol intake | Alcohol intake (g/day) | Alcohol (ID: 26030)[2] |

| | | |
|---|---|---|
| Smoking status | Never; Previous; Current; Unknown/Missing | Smoking status (ID: 20116)[1] |
| Physical activity | METs hr/week quintiles; Unknown/Missing | Duration of walks (ID: 874)[1]; Number of days/week walked 10+ minutes (ID: 864)[1]; Duration of moderate activity (ID: 894)[1]; Number of days/week of moderate physical activity 10+ minutes (ID: 884)[1]; Duration of vigorous activity (ID: 914)[1]; Number of days/week of vigorous physical activity 10+ minutes (ID: 904) [1] |
| Energy intake | Energy intake (kJ/day) | Energy (ID: 26002)[2] |
| Number of dietary assessments completed | Number of dietary assessments completed (ranging from 2-5) | Number of diet questionnaires completed (ID: 20077)[2] |
| Total protein intake | Total protein intake (g/day) | Diet by 24-hour recall (category ID: 100090)[2] |
| **Health status** | | |
| BMI | Healthy weight (18.5-24.99 kg/m$^2$); Overweight (25-29.99 kg/m$^2$); Obese (≥30 kg/m$^2$); Unknown/Missing | BMI (ID: 21001)[3] |
| Waist circumference | Waist circumference (cm) | Waist circumference (ID: 48)[3] |
| Multimorbidity | Number of pre-existing long-term conditions (0, 1, 2, ≥3) | Non-cancer illness diagnosed by nurse during verbal interview (ID: 20002)[4]; Cancer diagnosed by doctor (ID: 2453)[1] |
| Diabetes type | Type 1; Type 2; Unspecified | ICD10 codes: Diabetes (E10-E14); Medication for cholesterol, blood pressure or diabetes (men) (ID: 6177)[1]; Medication for cholesterol, blood pressure, diabetes, or take exogenous hormones (women) (ID: 6153)[1]; Non-cancer illness diagnosed by nurse during verbal interview (ID: 20002)[4] |
| Diabetes medication | No; Yes; Unknown/Missing | Medication for cholesterol, blood pressure or diabetes (men) (ID: 6177)[1]; Medication for cholesterol, blood pressure, diabetes, or take exogenous hormones (women) (ID: 6153)[1]; Treatment/medication code (ID: 20003)[4] |
| eGFR | eGFR (ml/min/1.73m$^2$) | CKD-EPI creatinine equation (4); Serum creatinine (ID: 30700)[3] |
| Nephrolithiasis | No; Yes | Non-cancer illness diagnosed by nurse during verbal interview (ID: 20002)[4] |
| Hypertension | No; Yes | Vascular/heart problems diagnosed by doctor (heart attack, angina, stroke) (ID: 6150)[1]; Medication for cholesterol, blood pressure or diabetes (men) (ID: 6177)[1]; Medication for cholesterol, blood pressure, diabetes, or take exogenous hormones (women) (ID: 6153)[1]; Non-cancer illness diagnosed by nurse during verbal interview (ID: 20002)[4] |

| | | |
|---|---|---|
| Systolic blood pressure | Systolic blood pressure (mmHg) | Systolic blood pressure, automated reading (ID: 4080)[3] |
| PRS (CKD) | Tertiles from low to high PRS for eGFR; Unknown/Missing | Genomics (category ID: 100314)[5] |
| **Blood biomarkers** | | |
| LDL cholesterol | LDL-direct (mmol/L) | LDL-direct (ID: 30780)[3] |
| HDL cholesterol | HDL (mmol/L) | HDL (ID: 30760)[3] |
| Triglycerides | Triglycerides (mmol/L) | Triglycerides (ID: 30870)[3] |
| Lipoprotein A | Lipoprotein A (nmol/L) | Lipoprotein A (ID:30790)[3] |
| Apolipoprotein A | Apolipoprotein A (g/L) | Apolipoprotein A (ID: 30630)[3] |
| Cystatin C | Cystatin C (mg/L) | Cystatin C (ID: 30720)[3] |
| Urate | Urate (umol/L) | Urate (ID: 30880)[3] |
| Creatinine | Creatinine (umol/L) | Creatinine (ID: 30700)[3] |
| C-reactive protein | C-reactive protein (mg/L) | C-reactive protein (ID: 30710)[3] |
| Albumin | Albumin (g/L) | Albumin (ID: 30600)[3] |

[1]Data collected at recruitment via touchscreen questionnaire (initial assessment visit (2006-2010)
[2]Data collected from 24-hr online Oxford WebQ dietary questionnaire (assessment centre (April 2009 to September 2010; on-line cycle 1 (February 2011 to April 2011); on-line cycle 2 (June 2011 to September 2011); on-line cycle 3 (October 2011 to December 2011); on-line cycle 4 (April 2012 to June 2012)
[3]Physical measurements (initial assessment visit (2006-2010)
[4]Data collected via verbal interview (initial assessment visit (2006-2010)
[5]Genomics data (using blood samples from initial assessment visit (2006-2010)
*Abbreviations: MET, metabolic equivalent task; BMI, body mass index; eGFR, estimated glomerular filtration rate; CKD, chronic kidney disease; PRS, polygenic risk score; LDL, low-density lipoprotein; HDL, high-density lipoprotein.*

**Table S2**: Genetic instruments for eGFR (n=161 SNPs)

| SNP | Chr | Pos_(b37) | Locus | EA | OA | EAF | Effect | SE | p-value |
|---|---|---|---|---|---|---|---|---|---|
| **rs74748843** | 1 | 10730910 | CASZ1 | T | C | 0.07 | -0.0048 | 8e-4 | 3.7e-9 |
| **rs10159261** | 1 | 15912987 | AGMAT | T | G | 0.36 | -0.0034 | 3e-4 | 4.8e-25 |
| **rs12061708** | 1 | 18809916 | KLHDC7A | A | G | 0.29 | -0.0026 | 3e-4 | 9.6e-14 |
| **rs659437** | 1 | 46037394 | AKR1A1 | T | C | 0.22 | -0.0027 | 4e-4 | 3.3e-12 |
| **rs11211257** | 1 | 46581933 | PIK3R3 | A | G | 0.82 | 0.0027 | 5e-4 | 2.3e-9 |
| **rs688540** | 1 | 48002447 | FOXD2 | A | G | 0.87 | -0.003 | 5e-4 | 3e-8 |
| **rs17413465** | 1 | 55718708 | MIR4422HG | A | C | 0.18 | 0.0025 | 4e-4 | 8.9e-9 |
| **rs1757915** | 1 | 56615809 | LINC01755 | A | G | 0.33 | 0.0021 | 3e-4 | 2.9e-10 |
| **rs7536433** | 1 | 78023173 | AK5 | T | C | 0.26 | 0.0021 | 4e-4 | 6.7e-9 |
| **rs679843** | 1 | 78707493 | MGC27382 | T | C | 0.33 | 0.0021 | 3e-4 | 5.2e-10 |
| **rs1887252** | 1 | 82957871 | LINC01362 | C | G | 0.62 | -0.0019 | 3e-4 | 2.9e-9 |
| **rs11166440** | 1 | 100808363 | CDC14A | A | G | 0.6 | 0.002 | 3e-4 | 1.8e-10 |
| **rs10857788** | 1 | 110012289 | SYPL2 | A | G | 0.7 | 0.003 | 4e-4 | 2e-16 |
| **rs12736457** | 1 | 113258293 | PPM1J | C | G | 0.87 | 0.0054 | 5e-4 | 1e-25 |
| **rs267738** | 1 | 150940625 | CERS2 | T | G | 0.8 | -0.0048 | 4e-4 | 1.20e-32 |
| **rs3845534** | 1 | 163738950 | LOC100422212 | A | G | 0.53 | -0.0019 | 3e-4 | 1.2e-9 |
| **rs4656220** | 1 | 170649277 | PRRX1 | T | C | 0.42 | 0.002 | 3e-4 | 3.3e-10 |
| **rs3795503** | 1 | 180905694 | KIAA1614 | T | C | 0.36 | 0.002 | 3e-4 | 9.8e-10 |
| **rs78329830** | 1 | 186769572 | PLA2G4A | A | G | 0.96 | -0.0054 | 9e-4 | 6.7e-9 |
| **rs3850625** | 1 | 201016296 | CACNA1S | A | G | 0.12 | 0.0046 | 5e-4 | 1.1e-18 |
| **rs75625374** | 1 | 208039431 | CD34 | C | G | 0.06 | 0.0045 | 7e-4 | 4.5e-10 |
| **rs7535253** | 1 | 214744893 | PTPN14 | T | C | 0.27 | 0.0021 | 4e-4 | 1.3e-9 |
| **rs61830291** | 1 | 221001142 | LINC01352 | A | C | 0.9 | -0.0036 | 6e-4 | 1.2e-9 |
| **rs417237** | 1 | 228532195 | OBSCN | T | G | 0.57 | 0.0018 | 3e-4 | 7.5e-9 |
| **rs2490391** | 1 | 243469669 | SDCCAG8 | A | C | 0.43 | -0.0024 | 3e-4 | 1.3e-14 |
| **rs3791221** | 2 | 226933 | SH3YL1 | A | G | 0.67 | 0.0022 | 3e-4 | 1.2e-11 |
| **rs2301343** | 2 | 40680149 | SLC8A1 | T | G | 0.76 | -0.0023 | 4e-4 | 4.1e-10 |
| **rs10865189** | 2 | 43433257 | ZFP36L2 | C | G | 0.51 | 0.0024 | 3e-4 | 3.3e-14 |
| **rs2971880** | 2 | 54885640 | SPTBN1 | A | T | 0.37 | -0.0024 | 3e-4 | 7.6e-15 |
| **rs10197255** | 2 | 67874553 | LINC01812 | A | T | 0.4 | 0.0018 | 3e-4 | 1.2e-8 |
| **rs11694902** | 2 | 121988884 | TFCP2L1 | A | G | 0.14 | 0.0041 | 5e-4 | 1.1e-16 |
| **rs7425436** | 2 | 148759656 | ORC4 | A | G | 0.65 | 0.0024 | 3e-4 | 5.2e-13 |
| **rs35472707** | 2 | 169995581 | LRP2 | T | C | 0.05 | -0.0073 | 8e-4 | 6.2e-19 |
| **rs187355703** | 2 | 176993583 | HOXD8 | C | G | 0.97 | 0.01 | 0.0011 | 1e-18 |
| **rs4666821** | 2 | 183077254 | PDE1A | T | G | 0.53 | 0.002 | 3e-4 | 2.5e-11 |
| **rs60980181** | 2 | 188168567 | CALCRL | A | T | 0.17 | -0.0027 | 4e-4 | 1.8e-10 |
| **rs1047891** | 2 | 211540507 | CPS1 | A | C | 0.29 | -0.0065 | 4e-4 | 1.2e-75 |
| **rs1548945** | 2 | 217665788 | TNP1 | T | C | 0.44 | 0.0036 | 3e-4 | 8.4e-31 |
| **rs1050816** | 2 | 220358198 | SPEG | T | C | 0.33 | 0.0026 | 3e-4 | 1.1e-15 |
| **rs35669853** | 2 | 227287718 | MIR5702 | A | G | 0.18 | 0.0024 | 4e-4 | 9.2e-9 |
| **rs795009** | 3 | 12208671 | SYN2 | T | G | 0.73 | 0.002 | 3e-4 | 7e-9 |
| **rs6778731** | 3 | 13947504 | WNT7A | T | C | 0.59 | -0.0017 | 3e-4 | 3.3e-8 |
| **rs6779998** | 3 | 30749965 | TGFBR2 | A | G | 0.52 | -0.0017 | 3e-4 | 1.6e-8 |
| **rs7651407** | 3 | 48443816 | PLXNB1 | T | C | 0.44 | 0.0025 | 4e-4 | 2.4e-11 |
| **rs4625** | 3 | 49572140 | DAG1 | A | G | 0.7 | -0.0023 | 4e-4 | 5.5e-11 |
| **rs2581820** | 3 | 53020544 | SFMBT1 | A | G | 0.29 | 0.0021 | 3e-4 | 7.9e-10 |

| | | | | | | | | | |
|---|---|---|---|---|---|---|---|---|---|
| **rs2289746** | 3 | 105455955 | CBLB | T | C | 0.41 | -0.0019 | 3e-4 | 2.5e-9 |
| **rs9868185** | 3 | 121657593 | SLC15A2 | A | G | 0.5 | 0.0026 | 3e-4 | 5e-17 |
| **rs10934754** | 3 | 125906237 | ALDH1L1-AS2 | T | C | 0.6 | 0.002 | 3e-4 | 1.3e-10 |
| **rs35320690** | 3 | 135932494 | MSL2 | T | C | 0.73 | -0.0025 | 4e-4 | 3e-11 |
| **rs9828976** | 3 | 136536835 | SLC35G2 | C | G | 0.76 | -0.0024 | 4e-4 | 1.9e-9 |
| **rs1397764** | 3 | 141750810 | TFDP2 | A | G | 0.27 | 0.0043 | 3e-4 | 2.5e-37 |
| **rs76272256** | 3 | 168888112 | MECOM | T | C | 0.24 | 0.0024 | 4e-4 | 4.9e-10 |
| **rs56065557** | 3 | 185354216 | SENP2 | C | G | 0.32 | -0.0029 | 3e-4 | 4.3e-18 |
| **rs11919484** | 3 | 186432839 | KNG1 | T | G | 0.32 | -0.0026 | 3e-4 | 5.8e-16 |
| **rs75501914** | 4 | 3449781 | HGFAC | A | G | 0.09 | 0.0039 | 6e-4 | 9e-11 |
| **rs3775932** | 4 | 10090930 | WDR1 | A | C | 0.51 | -0.0018 | 3e-4 | 2e-9 |
| **rs16874073** | 4 | 23743962 | PPARGC1A | T | C | 0.95 | -0.0045 | 7e-4 | 6.6e-11 |
| **rs12509595** | 4 | 81182554 | FGF5 | T | C | 0.7 | -0.0035 | 3e-4 | 6.4e-25 |
| **rs223471** | 4 | 103698786 | LOC102723704 | C | G | 0.34 | 0.0028 | 3e-4 | 5.9e-19 |
| **rs55929207** | 4 | 109703549 | ETNPPL | C | G | 0.48 | 0.0019 | 3e-4 | 3.4e-10 |
| **rs71606723** | 4 | 115498457 | UGT8 | A | T | 0.77 | 0.0025 | 4e-4 | 3.4e-12 |
| **rs1362800** | 5 | 39378115 | DAB2 | T | C | 0.38 | -0.0049 | 3e-4 | 5.8e-51 |
| **rs495237** | 5 | 39950266 | LINC00603 | T | G | 0.25 | 0.0027 | 3e-4 | 2e-14 |
| **rs11746506** | 5 | 44812566 | MRPS30 | T | C | 0.41 | 0.0017 | 3e-4 | 3e-8 |
| **rs12520984** | 5 | 52787358 | FST | C | G | 0.32 | 0.0019 | 3e-4 | 5.3e-9 |
| **rs79760705** | 5 | 53298716 | ARL15 | T | G | 0.11 | 0.0056 | 5e-4 | 6.5e-25 |
| **rs72759880** | 5 | 67750213 | PIK3R1 | T | G | 0.11 | -0.0056 | 5e-4 | 1.1e-26 |
| **rs3797537** | 5 | 78322650 | DMGDH | A | G | 0.73 | 0.0019 | 3e-4 | 2.9e-8 |
| **rs12777** | 5 | 131671662 | SLC22A4 | C | G | 0.96 | 0.005 | 9e-4 | 1.1e-8 |
| **rs12163971** | 5 | 132226669 | AFF4 | A | C | 0.16 | -0.0029 | 4e-4 | 1.7e-12 |
| **rs11743174** | 5 | 148524820 | ABLIM3 | T | C | 0.67 | 0.0019 | 3e-4 | 1.3e-8 |
| **rs3812036** | 5 | 176813404 | SLC34A1 | T | C | 0.26 | -0.0065 | 4e-4 | 2.4e-74 |
| **rs144100226** | 6 | 34180297 | HMGA1 | T | C | 0.04 | 0.0059 | 0.001 | 6e-9 |
| **rs77915916** | 6 | 43287722 | CRIP3 | A | T | 0.92 | 0.0046 | 6e-4 | 7.3e-14 |
| **rs720989** | 6 | 44765535 | SUPT3H | T | G | 0.79 | 0.0021 | 4e-4 | 1.8e-8 |
| **rs12212034** | 6 | 51492862 | PKHD1 | T | C | 0.37 | -0.0018 | 3e-4 | 1e-8 |
| **rs6458868** | 6 | 52630153 | GSTA2 | T | C | 0.67 | -0.002 | 3e-4 | 1.2e-9 |
| **rs3925003** | 6 | 55422618 | HMGCLL1 | T | C | 0.58 | -0.0018 | 3e-4 | 2e-9 |
| **rs72912510** | 6 | 90118764 | RRAGD | A | G | 0.2 | -0.0024 | 4e-4 | 6.4e-9 |
| **rs1857859** | 6 | 100894587 | SIM1 | A | G | 0.31 | 0.0019 | 3e-4 | 2.6e-8 |
| **rs7740107** | 6 | 130374461 | L3MBTL3 | A | T | 0.74 | 0.0027 | 4e-4 | 8.9e-13 |
| **rs9375818** | 6 | 131882078 | ARG1 | A | G | 0.25 | -0.0031 | 4e-4 | 6.1e-18 |
| **rs9397738** | 6 | 154986664 | SCAF8 | A | G | 0.84 | 0.0027 | 4e-4 | 3.5e-10 |
| **rs12207180** | 6 | 160633107 | SLC22A2 | A | T | 0.11 | -0.0085 | 5e-4 | 2.6e-63 |
| **rs6968554** | 7 | 17287106 | AHR | A | G | 0.43 | -0.0019 | 3e-4 | 1.1e-9 |
| **rs700753** | 7 | 46753684 | LOC730338 | C | G | 0.32 | 0.0031 | 3e-4 | 2.1e-20 |
| **rs55773927** | 7 | 65337902 | VKORC1L1 | T | C | 0.41 | 0.0019 | 3e-4 | 1.2e-8 |
| **rs801193** | 7 | 66030612 | GS1-124K5.11 | T | G | 0.58 | -0.002 | 3e-4 | 1.9e-9 |
| **rs41301394** | 7 | 75612803 | POR | T | C | 0.32 | 0.0023 | 3e-4 | 3.7e-12 |
| **rs6973656** | 7 | 77422583 | TMEM60 | A | G | 0.64 | 0.0035 | 3e-4 | 5.70e-28 |
| **rs3757387** | 7 | 128576086 | IRF5 | T | C | 0.59 | 0.003 | 3e-4 | 7e-20 |
| **rs62491533** | 7 | 129564134 | UBE2H | T | C | 0.81 | -0.0027 | 4e-4 | 1.1e-11 |

| | | | | | | | | | |
|---|---|---|---|---|---|---|---|---|---|
| **rs1533059** | 8 | 8684953 | MFHAS1 | A | G | 0.51 | 0.0025 | 3e-4 | 1.2e-14 |
| **rs10098664** | 8 | 11417493 | BLK | T | C | 0.49 | -0.0021 | 3e-4 | 6.1e-10 |
| **rs2976178** | 8 | 87332552 | WWP1 | C | G | 0.67 | -0.0025 | 3e-4 | 7.8e-14 |
| **rs2954017** | 8 | 126476873 | TRIB1 | T | C | 0.46 | 0.0024 | 3e-4 | 1.7e-12 |
| **rs12377027** | 9 | 20554583 | MLLT3 | A | G | 0.82 | -0.0026 | 5e-4 | 2.9e-8 |
| **rs13287724** | 9 | 33169034 | B4GALT1-AS1 | A | T | 0.89 | -0.003 | 6e-4 | 4.7e-8 |
| **rs2039424** | 9 | 71432174 | PIP5K1B | A | G | 0.64 | 0.0044 | 3e-4 | 2.1e-44 |
| **rs1321917** | 9 | 119324929 | ASTN2 | C | G | 0.44 | -0.0023 | 3e-4 | 1.4e-13 |
| **rs7024579** | 9 | 139100413 | QSOX2 | T | C | 0.29 | 0.0023 | 4e-4 | 8.2e-11 |
| **rs80282103** | 10 | 899071 | LARP4B | A | T | 0.91 | 0.0078 | 6e-4 | 1.2e-44 |
| **rs6481598** | 10 | 29781798 | SVIL | C | G | 0.79 | 0.0024 | 4e-4 | 1.5e-9 |
| **rs10821905** | 10 | 52646093 | A1CF | A | G | 0.18 | 0.0037 | 4e-4 | 9.4e-19 |
| **rs10821944** | 10 | 63785089 | ARID5B | T | G | 0.7 | 0.002 | 3e-4 | 3.9e-9 |
| **rs7475348** | 10 | 69965177 | MYPN | T | C | 0.46 | 0.0031 | 3e-4 | 1.2e-22 |
| **rs816850** | 10 | 79252446 | KCNMA1 | C | G | 0.25 | -0.002 | 4e-4 | 7.4e-9 |
| **rs7095954** | 10 | 82209232 | TSPAN14 | A | T | 0.45 | -0.0018 | 3e-4 | 3.7e-8 |
| **rs4918943** | 10 | 97278922 | SORBS1 | A | G | 0.22 | -0.0022 | 4e-4 | 9.2e-9 |
| **rs284859** | 10 | 104573017 | WBP1L | T | G | 0.21 | 0.0026 | 4e-4 | 5e-12 |
| **rs1055256** | 10 | 126446592 | EEF1AKMT2 | A | G | 0.42 | 0.0025 | 3e-4 | 3.6e-16 |
| **rs11564722** | 11 | 2178330 | INS-IGF2 | T | C | 0.31 | 0.0033 | 4e-4 | 2.1e-20 |
| **rs63934** | 11 | 2789062 | KCNQ1 | A | G | 0.83 | 0.0041 | 4e-4 | 3.90e-23 |
| **rs61897431** | 11 | 47427667 | SLC39A13 | T | C | 0.65 | 0.0029 | 4e-4 | 6.3e-16 |
| **rs948493** | 11 | 65552154 | MIR1234 | T | C | 0.33 | -0.0033 | 3e-4 | 2e-24 |
| **rs6589750** | 11 | 119326726 | USP2-AS1 | A | G | 0.63 | 0.002 | 3e-4 | 1.5e-9 |
| **rs117113238** | 12 | 12209203 | BCL2L14 | A | G | 0.09 | 0.0039 | 6e-4 | 8.6e-11 |
| **rs41284816** | 13 | 50655989 | DLEU2 | T | G | 0.03 | -0.0078 | 0.0012 | 1.7e-10 |
| **rs72683923** | 14 | 50735947 | L2HGDH | T | C | 0.98 | -0.0074 | 0.0013 | 3.4e-8 |
| **rs1028455** | 14 | 88829975 | SPATA7 | A | T | 0.33 | 0.002 | 3e-4 | 4.8e-10 |
| **rs17184313** | 14 | 93102251 | RIN3 | T | C | 0.17 | -0.0029 | 5e-4 | 2e-10 |
| **rs61993680** | 14 | 100752644 | SLC25A29 | A | C | 0.61 | -0.0019 | 3e-4 | 1.5e-8 |
| **rs12913015** | 15 | 39305443 | C15orf54 | T | C | 0.41 | 0.0027 | 3e-4 | 2.3e-17 |
| **rs1994887** | 15 | 57793765 | CGNL1 | A | C | 0.26 | -0.002 | 4e-4 | 1.6e-8 |
| **rs956006** | 15 | 62808539 | MGC15885 | T | C | 0.33 | 0.0019 | 3e-4 | 4.4e-9 |
| **rs11071939** | 15 | 67463391 | SMAD3 | T | C | 0.93 | -0.0039 | 6e-4 | 4.9e-10 |
| **rs4886755** | 15 | 76298132 | NRG4 | A | G | 0.5 | 0.0041 | 3e-4 | 2e-39 |
| **rs17507300** | 15 | 83722059 | BTBD1 | A | G | 0.83 | 0.0024 | 4e-4 | 1e-8 |
| **rs7169629** | 15 | 85191274 | WDR73 | C | G | 0.48 | 0.0018 | 3e-4 | 1.6e-8 |
| **rs59646751** | 15 | 99276521 | IGF1R | T | G | 0.3 | -0.0023 | 3e-4 | 3.1e-12 |
| **rs438339** | 16 | 2003425 | RPL3L | T | C | 0.88 | 0.0035 | 6e-4 | 5e-8 |
| **rs77924615** | 16 | 20392332 | PDILT | A | G | 0.2 | 0.0098 | 4e-4 | 1.5e-138 |
| **rs9932625** | 16 | 51735746 | LINC01571 | A | G | 0.26 | -0.003 | 3e-4 | 2.2e-17 |
| **rs7203398** | 16 | 53189672 | CHD9 | A | C | 0.74 | 0.0025 | 3e-4 | 4.7e-13 |
| **rs62050038** | 16 | 69802865 | WWP2 | A | T | 0.83 | 0.0028 | 4e-4 | 1.3e-11 |
| **rs62053077** | 16 | 71643669 | MARVELD3 | T | G | 0.43 | -0.0021 | 4e-4 | 3.7e-9 |
| **rs1858800** | 16 | 73024276 | ZFHX3 | T | C | 0.32 | 0.002 | 3e-4 | 2.1e-9 |
| **rs28581385** | 16 | 79942679 | LINC01229 | A | T | 0.84 | -0.0028 | 4e-4 | 1.4e-11 |
| **rs2440165** | 17 | 19428719 | SLC47A1 | T | C | 0.64 | 0.004 | 3e-4 | 1.70e-31 |
| **rs35662455** | 17 | 56755223 | TEX14 | C | G | 0.89 | 0.003 | 5e-4 | 3.9e-8 |
| **rs883541** | 17 | 66449122 | PRKAR1A | A | G | 0.72 | -0.0022 | 3e-4 | 2.7e-10 |

| | | | | | | | | | |
|---|---|---|---|---|---|---|---|---|---|
| **rs1719934** | 18 | 5585158 | EPB41L3 | A | G | 0.58 | 0.0026 | 3e-4 | 2.6e-17 |
| **rs16942751** | 18 | 24393213 | AQP4 | A | C | 0.18 | -0.0029 | 5e-4 | 2.2e-9 |
| **rs2974751** | 19 | 13053034 | CALR | A | C | 0.38 | 0.0018 | 3e-4 | 4.4e-8 |
| **rs8101667** | 19 | 33402419 | CEP89 | T | C | 0.39 | 0.0044 | 3e-4 | 9.30e-44 |
| **rs34647824** | 19 | 50138143 | RRAS | A | C | 0.74 | -0.0021 | 4e-4 | 4e-8 |
| **rs62187537** | 20 | 1333060 | FKBP1A-SDCBP2 | T | C | 0.07 | 0.0039 | 7e-4 | 9.2e-9 |
| **rs1041606** | 20 | 14677788 | MACROD2 | T | C | 0.23 | -0.0021 | 4e-4 | 2.5e-8 |
| **rs6087579** | 20 | 32985155 | ITCH | A | G | 0.48 | -0.0028 | 3e-4 | 1.4e-19 |
| **rs2235826** | 20 | 56143169 | PCK1 | A | T | 0.79 | -0.003 | 4e-4 | 6.8e-15 |
| **rs1407040** | 20 | 57472174 | GNAS | T | C | 0.68 | 0.0018 | 3e-4 | 1.4e-8 |
| **rs35636653** | 20 | 60858758 | OSBPL2 | T | C | 0.34 | 0.0022 | 3e-4 | 2.3e-11 |
| **rs4408777** | 20 | 62706105 | RGS19 | A | G | 0.51 | -0.0021 | 3e-4 | 5.4e-11 |
| **rs2823139** | 21 | 16576783 | NRIP1 | A | G | 0.33 | -0.0026 | 3e-4 | 5.2e-16 |
| **rs2834317** | 21 | 35356706 | LOC101928126 | A | G | 0.14 | -0.0035 | 5e-4 | 4.3e-14 |
| **rs2244237** | 21 | 37818141 | CLDN14 | T | G | 0.22 | 0.0027 | 4e-4 | 6.3e-11 |
| **rs80576** | 22 | 36539804 | APOL3 | A | G | 0.16 | -0.0028 | 5e-4 | 1.3e-9 |
| **rs4820324** | 22 | 38599857 | MAFF | C | G | 0.59 | -0.0023 | 3e-4 | 5.1e-14 |
| **rs112880707** | 22 | 40884662 | MKL1 | T | C | 0.17 | 0.0052 | 5e-4 | 4.90e-31 |
| **rs738527** | 22 | 43112961 | A4GALT | T | C | 0.29 | 0.0032 | 3e-4 | 4.2e-21 |

*Abbreviations: SNP, single nucleotide polymorphism; Chr, chromosome; Pos_(b37), position and human genome reference build; EA, effect allele; OA, other allele; EAF, effect allele frequency; SE, standard error.*

**Table S3.** Baseline characteristics across quartiles (Q) of the unhealthful plant-based diet index in the UK Biobank (n=7,747)

| | Participants, No. (%)[1] | | | | |
|---|---|---|---|---|---|
| **Characteristics across uPDI** | **Q1** | **Q2** | **Q3** | **Q4** | **Whole sample** |
| **Number of participants** | 1,983 (25.6) | 2,295 (29.6) | 1,790 (23.1) | 1,679 (21.7) | 7,747 (100.0) |
| **CKD cases** | 232 (11.7) | 308 (13.4) | 248 (13.9) | 242 (14.4) | 1,030 (13.3) |
| **Deaths** | 205 (10.3) | 263 (11.5) | 210 (11.7) | 207 (12.3) | 885 (11.4) |
| **Unhealthful plant-based diet index, mean (SD)** | 47.4 (3.1) | 53.7 (1.5) | 58.0 (1.2) | 63.6 (2.9) | 55.2 (6.2) |
| **Sex-Female** | 760 (38.3) | 885 (38.9) | 697 (38.9) | 686 (40.9) | 3,038 (39.2) |
| **Age at recruitment (years), mean (SD)** | 59.9 (6.8) | 59.4 (6.9) | 58.4 (7.4) | 57.2 (7.6) | 58.8 (7.2) |
| **BMI (kg/m$^2$), mean (SD)** | 30.2 (5.7) | 30.4 (5.7) | 31.0 (5.9) | 31.8 (5.9) | 30.8 (5.9) |
| **Waist circumference (cm), mean (SD)** | 99.7 (14.8) | 99.9 (14.4) | 101.7 (14.8) | 103.6 (14.1) | 101.1 (14.6) |
| **Energy intake (kJ/day), mean (SD)** | 8809.2 (2090.6) | 8386.1 (2163.3) | 8149.7 (2193.4) | 7990.3 (2285.3) | 8354.0 (2199.7) |
| **Physical activity (MET-h/wk), mean (SD)** | 29.5 (37.8) | 29.2 (38.5) | 26.6 (37.9) | 25.9 (39.4) | 28.0 (38.4) |
| **Ethnicity** | | | | | |
| Asian | 107 (5.4) | 97 (4.2) | 60 (3.4) | 53 (3.2) | 317 (4.1) |
| Black | 27 (1.4) | 27 (1.2) | 24 (1.3) | 23 (1.4) | 101 (1.3) |
| Multiple | 68 (3.4) | 76 (3.3) | 67 (3.7) | 71 (4.2) | 282 (3.6) |
| White | 1,745 (88.0) | 2,053 (89.5) | 1,609 (89.9) | 1,503 (89.5) | 6,910 (89.2) |
| Other/missing[2] | 36 (1.8) | 42 (1.8) | 30 (1.7) | 29 (1.7) | 137 (1.8) |
| **Education** | | | | | |
| Low | 451 (22.7) | 529 (23.1) | 433 (24.2) | 436 (26.0) | 1,849 (23.9) |
| Medium | 355 (17.9) | 396 (17.3) | 351 (19.6) | 308 (18.3) | 1,410 (18.2) |
| High | 956 (48.2) | 1,035 (45.1) | 727 (40.6) | 598 (35.6) | 3,316 (42.8) |
| Missing | 221 (11.1) | 335 (14.6) | 279 (15.6) | 337 (20.1) | 1,172 (15.1) |
| **Smoking status** | | | | | |
| Never | 858 (43.3) | 1,034 (45.1) | 826 (46.2) | 840 (50.0) | 3,558 (45.9) |
| Previous | 970 (48.9) | 1,072 (46.7) | 796 (44.5) | 662 (39.4) | 3,500 (45.2) |
| Current | 144 (7.3) | 176 (7.7) | 161 (9.0) | 170 (10.1) | 651 (8.4) |
| Missing | 11 (1.0) | 13 (1.0) | 7 (0.4) | 7 (0.4) | 38 (0.5) |
| **Alcohol intake (g/day), mean (SD)** | 15.2 (20.9) | 14.7 (21.3) | 16.2 (25.1) | 14.0 (24.7) | 15.0 (22.9) |
| **Diabetes type** | | | | | |
| Type 1 diabetes | 341 (17.2) | 394 (17.2) | 271 (15.1) | 247 (14.7) | 1,253 (16.2) |
| Type 2 diabetes | 1,306 (65.9) | 1,531 (66.7) | 1,228 (68.6) | 1,150 (68.5) | 5,215 (67.3) |
| Unspecified | 336 (16.9) | 370 (16.1) | 291 (16.3) | 282 (16.8) | 1,279 (16.5) |
| **Hypertension** | 1,132 (57.1) | 1,341 (58.4) | 1,094 (61.1) | 985 (58.7) | 4,552 (58.8) |
| **HbA1c (mmol/mol), mean (SD)** | 50.3 (12.7) | 51.3 (13.6) | 51.4 (13.5) | 51.3 (13.7) | 51.1 (13.4) |
| **SBP (mm Hg), mean (SD)** | 137.0 (17.1) | 137.0 (16.5) | 135.4 (16.3) | 135.7 (17.2) | 136.3 (16.8) |
| **DBP (mm Hg), mean (SD)** | 81.8 (10.5) | 82.4 (10.8) | 83.1 (11.4) | 83.2 (9.9) | 82.6 (10.6) |
| **Multimorbidity** | | | | | |
| 0 LTCs | 475 (24.0) | 508 (22.1) | 389 (21.7) | 395 (23.5) | 1,767 (22.8) |
| 1 LTC | 735 (37.1) | 824 (35.9) | 642 (35.9) | 540 (32.2) | 2,741 (35.4) |
| 2 LTCs | 472 (23.8) | 555 (24.2) | 455 (25.4) | 409 (24.4) | 1,891 (24.4) |
| ≥3 LTCs | 301 (15.2) | 408 (17.8) | 304 (17.0) | 335 (20.0) | 1,348 (17.4) |
| **eGFR (mL/min/1.73 m2), mean (SD),** | 92.0 (11.8) | 91.6 (12.3) | 92.5 (12.9) | 93.2 (13.2) | 92.3 (12.5) |
| **Protein intake (g/day), mean (SD)** | 93.3 (24.0) | 83.9 (23.1) | 77.5 (22.5) | 71.4 (24.1) | 82.1 (24.7) |
| **hPDI food item intake, mean (SD) portions/day[3]** | | | | | |
| **Healthy plant food** | | | | | |
| Whole grains | 3.2 (1.8) | 2.4 (1.8) | 1.9 (1.6) | 1.2 (1.4) | 2.2 (1.8) |
| Fruit | 3.1 (1.9) | 2.5 (1.7) | 2.0 (1.6) | 1.5 (1.5) | 2.3 (1.8) |
| Vegetables | 3.5 (2.6) | 2.4 (2.0) | 1.8 (1.8) | 1.3 (1.6) | 2.3 (2.2) |
| Nuts | 0.3 (0.5) | 0.2 (0.4) | 0.1 (0.3) | 0.0 (0.2) | 0.1 (0.4) |
| Legumes | 0.6 (0.7) | 0.4 (0.6) | 0.3 (0.5) | 0.2 (0.4) | 0.4 (0.6) |
| Tea and coffee | 4.9 (1.8) | 4.4 (1.8) | 4.0 (1.8) | 3.4 (1.8) | 4.2 (1.9) |
| **Unhealthy plant food** | | | | | |

| | | | | | |
|---|---|---|---|---|---|
| Refined grains | 0.7 (1.0) | 1.1 (1.2) | 1.4 (1.4) | 2.0 (1.6) | 1.2 (1.4) |
| Potatoes | 0.7 (0.6) | 0.7 (0.6) | 0.8 (0.7) | 0.9 (0.8) | 0.8 (0.7) |
| Sugar-sweetened beverages | 0.4 (0.8) | 0.6 (0.9) | 0.8 (1.1) | 1.3 (1.4) | 0.7 (1.1) |
| Fruit juices | 0.2 (0.4) | 0.3 (0.5) | 0.4 (0.6) | 0.5 (0.7) | 0.3 (0.6) |
| Sweets and desserts | 0.8 (1.0) | 1.2 (1.2) | 1.4 (1.4) | 1.9 (1.6) | 1.3 (1.3) |
| **Animal-derived food** | | | | | |
| Animal fat | 1.0 (1.4) | 0.8 (1.3) | 0.7 (1.2) | 0.5 (1.0) | 0.8 (1.3) |
| Dairy | 1.3 (0.9) | 1.0 (0.9) | 0.9 (0.9) | 0.7 (0.8) | 1.0 (0.9) |
| Eggs | 0.5 (0.7) | 0.3 (0.6) | 0.3 (0.5) | 0.2 (0.4) | 0.3 (0.6) |
| Fish or seafood | 0.4 (0.5) | 0.3 (0.5) | 0.3 (0.5) | 0.2 (0.5) | 0.3 (0.5) |
| Meat | 1.4 (1.1) | 1.3 (1.0) | 1.3 (1.1) | 1.2 (1.1) | 1.3 (1.1) |
| Miscellaneous animal-based foods | 0.1 (0.4) | 0.1 (0.4) | 0.1 (0.3) | 0.1 (0.3) | 0.1 (0.3) |

[1]Relative frequencies (%) include missing values which may not equate to 100%.

[2]Other includes any race or ethnic group not otherwise specified.

[3]Portion sizes were specified as a "serving" in the Oxford WebQ tool.

*Abbreviations: Q, quartile; uPDI, unhealthful plant-based diet index; BMI, body mass index; MET, metabolic equivalent task; SD, standard deviation; SBP, systolic blood pressure; DBP, diastolic blood pressure; LTC, long term condition; eGFR, estimated glomerular filtration rate.*

**Table S4.** Key nutrient intakes across quartiles (Q) of healthful plant-based diet index (N=7,747)

| **Key nutrient intakes** | **Mean (SD)** | | | |
|---|---|---|---|---|
| **Healthful plant-based diet index quartiles** | **Q1** | **Q2** | **Q3** | **Q4** |
| Participants, No. (%) | 1,901 (24.5) | 1,829 (23.6) | 1,836 (23.7) | 2,181 (28.2) |
| Healthful plant-based diet index | 49.4 (3.2) | 55.2 (1.2) | 59.0 (1.2) | 64.7 (3.0) |
| Energy, kJ/day | 9312.6 (2088.0) | 8481.2 (2134.0) | 8089.8 (2124.8) | 7634.2 (2093.3) |
| Calcium, mg/day | 1014.2 (327.6) | 955.4 (328.8) | 935.5 (316.5) | 934.7 (325.5) |
| Protein, g/day | 88.3 (25.0) | 82.5 (24.4) | 80.4 (24.2) | 77.8 (24.0) |
| Fibre, g/day | 15.5 (5.6) | 16.7 (6.0) | 18.2 (6.3) | 20.9 (6.9) |
| Total fat, g/day | 84.3 (25.6) | 71.9 (24.9) | 67.4 (24.6) | 61.5 (24.9) |
| Saturated fat, g/day | 32.4 (11.5) | 27.0 (10.6) | 24.4 (10.0) | 20.7 (9.4) |
| Cholesterol, mg/day | 325.1 (183.2) | 263.9 (170.6) | 239.9 (161.1) | 196.1 (138.7) |
| Iodine, µg/day | 222.7 (112.6) | 213.4 (110.4) | 200.6 (94.6) | 190.5 (95.9) |
| Vitamin B12, µg/day | 6.8 (4.0) | 6.4 (3.6) | 6.2 (3.6) | 5.7 (3.1) |
| Vitamin D, µg/day | 4.2 (2.9) | 3.7 (2.9) | 3.6 (3.0) | 3.2 (2.8) |

*Abbreviations: SD, standard deviation; Q, quartile.*

**Table S5.** Key nutrient intakes across quartiles (Q) of unhealthful plant-based diet index (N=7,747)

| **Key nutrient intakes** | **Mean (SD)** | | | |
|---|---|---|---|---|
| **Unhealthful plant-based diet index quartiles** | **Q1** | **Q2** | **Q3** | **Q4** |
| Participants, No. (%) | 1,983 (25.6) | 2,295 (29.6) | 1,790 (23.1) | 1,679 (21.7) |
| Unhealthful plant-based diet index | 47.4 (3.1) | 53.7 (1.5) | 58.0 (1.2) | 63.6 (2.9) |
| Energy, kJ/day | 8809.2 (2090.6) | 8386.1 (2163.3) | 8149.7 (2193.4) | 7990.3 (2285.3) |
| Calcium, mg/day | 1080.6 (327.8) | 979.3 (308.5) | 908.4 (314.1) | 842.9 (308.1) |
| Protein, g/day | 93.3 (24.0) | 83.9 (23.1) | 77.5 (22.5) | 71.4 (24.1) |
| Fibre, g/day | 21.9 (6.6) | 18.5 (5.9) | 16.4 (5.7) | 14.2 (5.5) |
| Total fat, g/day | 77.4 (26.2) | 71.2 (26.1) | 68.1 (26.4) | 66.0 (25.5) |
| Saturated fat, g/day | 27.4 (11.2) | 26.1 (11.1) | 25.2 (11.5) | 24.6 (10.9) |
| Cholesterol, mg/day | 318.2 (199.5) | 258.8 (158.7) | 231.0 (153.4) | 196.7 (134.7) |
| Iodine, µg/day | 238.2 (111.7) | 211.7 (96.6) | 196.1 (101.2) | 171.7 (95.6) |
| Vitamin B12, µg/day | 7.4 (3.7) | 6.4 (3.3) | 5.9 (3.4) | 5.1 (3.5) |
| Vitamin D, µg/day | 4.5 (3.4) | 3.7 (2.9) | 3.3 (2.6) | 2.9 (2.4) |

*Abbreviations: SD, standard deviation; Q, quartile.*

**Table S6.** Healthful plant-based diet score and incident chronic kidney disease stratified by UK Biobank population subgroups

| | Cases/ total | hPDI (10-point increments) | P-trend | P- interaction |
|---|---|---|---|---|
| **Age, years** | | | | |
| HR (95% CI)[1] | | | | 0.32 |
| <60 | 319/3,515 | 0.84 (0.69-1.03) | 0.09 | |
| ≥60 | 711/4,232 | 0.83 (0.73-0.95) | 0.005 | |
| **Sex** | | | | |
| HR (95% CI)[1] | | | | 0.48 |
| Male | 679/4,709 | 0.88 (0.77-1.01) | 0.06 | |
| Female | 351/3,038 | 0.77 (0.63-0.94) | 0.01 | |
| **BMI, kg/m$^2$** | | | | |
| HR (95% CI)[1] | | | | 0.87 |
| <25.0 | 92/1,100 | 0.70 (0.47-1.03) | 0.07 | |
| ≥ 25.0 | 927/6,621 | 0.86 (0.77-0.97) | 0.01 | |
| **Smoking status** | | | | |
| HR (95% CI)[1] | | | | 0.60 |
| Never | 397/ 3,558 | 0.85 (0.71-1.01) | 0.07 | |
| Ever | 628/4,151 | 0.83 (0.72-0.96) | 0.01 | |
| **Alcohol intake, g/day** | | | | |
| HR (95% CI)[1] | | | | 0.62 |
| Low | 454/ 3,020 | 0.84 (0.71-0.98) | 0.03 | |
| Moderate | 294/2,143 | 0.83 (0.67-1.04) | 0.10 | |
| High | 282/2,584 | 0.82 (0.66-1.02) | 0.07 | |
| **Total protein, g/day** | | | | |
| HR (95% CI)[1] | | | | 0.75 |
| <median | 526/3,867 | 0.92 (0.78-1.07) | 0.27 | |
| ≥median | 504/3,880 | 0.79 (0.68-0.92) | 0.003 | |
| **History of hypertension** | | | | |
| HR (95% CI)[1] | | | | 0.69 |
| No | 317/3,195 | 0.76 (0.62-0.93) | 0.008 | |
| Yes | 713/4,552 | 0.88 (0.77-1.00) | 0.06 | |
| **Diabetes type** | | | | |
| HR (95% CI)[1] | | | | 0.27 |
| Type 1 | 187/1,253 | 0.64 (0.49-0.85) | 0.002 | |
| Type 2 | 768/5,215 | 0.88 (0.78-1.00) | 0.06 | |

[1]Hazard Ratios with 95% Confidence Intervals (CI), adjusted for sex (excluding subgroup analysis), BMI (excluding subgroup analysis), waist circumference, ethnicity, physical activity, smoking status (excluding subgroup analysis), education, alcohol intake (excluding subgroup analysis), energy intake, multimorbidity index, Townsend deprivation index, type of diabetes (excluding subgroup analysis), diabetes medication use, number of completed dietary assessments, and total protein intake (excluding subgroup analysis); stratified by age (3-year categories) (excluding subgroup analysis) and region.

Heterogeneity was tested by comparing two models – one without an interaction term between subgroup of interest and hPDI (categorical), with a model that included an interaction term. The likelihood ratio test was used to produce P-interaction values.

*Abbreviations: Q, quartile; hPDI, healthful plant-based diet index; BMI, Body Mass Index; HR, hazard ratio; CI, confidence interval.*

**Table S7.** Sensitivity analyses showing hazard ratios (95% confidence intervals) across sex-specific healthful vs unhealthful plant-based diet index quartiles (Q), further adjusting for eGFR, laboratory measurements (LDL-C, HDL-C), lipoprotein(a), C-reactive protein, albumin) and nephrolithiasis, for participants who completed 1 or more dietary assessments and the associated risk of chronic kidney disease (n=5,405)

| **hPDI** | Q1 | Q2 | Q3 | Q4 | P-trend |
|---|---|---|---|---|---|
| Cases/total | 208/1,307 | 166/1,253 | 182/1,326 | 163/1,519 | |
| HR (95% CI)[1] | 1.00[2] | 0.78 (0.63-0.97) | 0.89 (0.72-1.10) | 0.75 (0.60-0.94) | 0.005 |
| **uPDI** | Q1 | Q2 | Q3 | Q4 | P-trend |
| Cases/total | 164/1,403 | 216/1,610 | 167/1,213 | 172/1,179 | |
| HR (95% CI)[1] | 1.00[2] | 1.19 (0.97-1.47) | 1.16 (0.93-1.46) | 1.29 (1.02-1.63) | 0.23 |

[1]Hazard Ratios with 95% Confidence Intervals (CI), adjusted for sex, BMI, waist circumference, ethnicity, physical activity, smoking status, education, alcohol intake, energy intake, multimorbidity index, Townsend deprivation index, type of diabetes, diabetes medication use, number of completed dietary assessments, total protein intake, eGFR, LDL-C, HDL-C, Lipoprotein(a), CRP, albumin and nephrolithiasis; stratified by age (3-year categories) and region.
P-trend is for linear trend.
[2]Reference categories.
*Abbreviations: Q, quartile; hPDI, healthful plant-based diet index; uPDI, unhealthful plant-based diet index; BMI, Body Mass Index; HR, hazard ratio; CI, confidence interval; eGFR, estimated glomerular filtration rate; LDL-C, low-density lipoprotein cholesterol; HDL, high-density lipoprotein cholesterol; CRP, c-reactive protein.*

**Table S8.** Sensitivity analyses showing hazard ratios (95% confidence intervals) across sex-specific healthful vs unhealthful plant-based diet index quartiles (Q), further adjusting for genetic susceptibility of kidney diseases, for participants who completed 1 or more dietary assessments and the associated risk of chronic kidney disease (n=7,747)

| **hPDI** | Q1 | Q2 | Q3 | Q4 | P-trend |
|---|---|---|---|---|---|
| Cases/total | 287/1,901 | 248/1,829 | 256/1,836 | 239/2,181 | |
| HR (95% CI)[1] | 1.00[2] | 0.88 (0.74-1.06) | 0.97 (0.81-1.16) | 0.76 (0.63-0.92) | 0.002 |
| **uPDI** | Q1 | Q2 | Q3 | Q4 | P-trend |
| Cases/total | 232/1,983 | 308/2,295 | 248/1,790 | 242/1,679 | |
| HR (95% CI)[1] | 1.00[2] | 1.20 (1.01-1.43) | 1.20 (0.99-1.45) | 1.35 (1.10- 1.65) | 0.007 |

[1]Hazard Ratios with 95% Confidence Intervals (CI), adjusted for sex, BMI, waist circumference, ethnicity, physical activity, smoking status, education, alcohol intake, energy intake, multimorbidity index, Townsend deprivation index, type of diabetes, diabetes medication use, number of completed dietary assessments, total protein intake, and PRS (eGFR); stratified by age (3-year categories) and region.

P-trend is for linear trend.

[2]Reference categories.

*Abbreviations: Q, quartile; hPDI, healthful plant-based diet index; uPDI, unhealthful plant-based diet index; BMI, Body Mass Index; HR, hazard ratio; CI, confidence interval; eGFR, estimated glomerular filtration rate; PRS, polygenic risk score*

**Table S9.** Sensitivity analyses showing hazard ratios (95% confidence intervals) across sex-specific healthful vs unhealthful plant-based diet index quartiles (Q), removing the first 2 years of follow-up for participants who completed 1 or more dietary assessments and the associated risk of chronic kidney disease (n=7,574)

| **hPDI** | Q1 | Q2 | Q3 | Q4 | P-trend |
|---|---|---|---|---|---|
| Cases/total | 265/1,855 | 238/1,792 | 235/1,788 | 228/2,139 | |
| HR (95% CI) | | | | | |
| Model 1 | 1.00[1] | 0.84 (0.70-1.00) | 0.83 (0.69-0.99) | 0.65 (0.54-0.78) | <0.001 |
| Model 2 | 1.00[1] | 0.91 (0.76-1.10) | 0.94 (0.78-1.13) | 0.77 (0.63-0.93) | 0.004 |
| Model 3 | 1.00[1] | 0.92 (0.77-1.10) | 0.95 (0.79-1.14) | 0.78 (0.64-0.94) | 0.006 |
| **uPDI** | Q1 | Q2 | Q3 | Q4 | P-trend |
| Cases/total | 218/1,938 | 293/2,250 | 229/1,752 | 226/1,634 | |
| HR (95% CI) | | | | | |
| Model 1 | 1.00[1] | 1.23 (1.03-1.47) | 1.28 (1.06-1.54) | 1.51 (1.25-1.83) | <0.001 |
| Model 2 | 1.00[1] | 1.23 (1.03-1.47) | 1.20 (0.99-1.46) | 1.39 (1.14- 1.69) | 0.002 |
| Model 3 | 1.00[1] | 1.21 (1.01-1.45) | 1.18 (0.97-1.43) | 1.35 (1.10- 1.66) | 0.01 |

Model 1 adjusted for sex and education; stratified by age (3-year categories) and region.
Model 2: Model 1 plus BMI, waist circumference, ethnicity, physical activity, smoking status, alcohol intake, energy intake, multimorbidity index, Townsend depriation index, type of diabetes, diabetes medication use and number of completed dietary assessments.
Model 3: Model 2 plus total protein intake.
P-trend is for linear trend.
[1]Reference categories.
*Abbreviations: Q, quartile; hPDI, healthful plant-based diet index; uPDI, unhealthful plant-based diet index; BMI, Body Mass Index; HR, hazard ratio; CI, confidence interval.*

**Table S10.** Sensitivity analyses showing hazard ratios (95% confidence intervals) across sex-specific healthful vs unhealthful plant-based diet index quartiles (Q), excluding sugar-sweetened beverages for participants who completed 1 or more dietary assessments and the associated risk of chronic kidney disease (n=7,747)

| **hPDI** | Q1 | Q2 | Q3 | Q4 | P-trend |
|---|---|---|---|---|---|
| Cases/total | 305/2,069 | 270/1,996 | 254/1,933 | 201/1,779 | |
| HR (95% CI)[1] | 1.00[2] | 0.92 (0.78-1.09) | 0.93 (0.78-1.11) | 0.77 (0.64-0.94) | 0.009 |
| **uPDI** | Q1 | Q2 | Q3 | Q4 | P-trend |
| Cases/total | 251/2,114 | 253/1,930 | 276/1,911 | 250/1,792 | |
| HR (95% CI)[1] | 1.00[2] | 1.14 (0.96-1.37) | 1.30 (1.09-1.56) | 1.24 (1.02- 1.50) | 0.02 |

[1]Hazard Ratios with 95% Confidence Intervals (CI), adjusted for sex, BMI, waist circumference, ethnicity, physical activity, smoking status, education, alcohol intake, energy intake, multimorbidity index, Townsend deprivation index, type of diabetes, diabetes medication use, number of completed dietary assessments, and total protein intake; stratified by age (3-year categories) and region.

P-trend is for linear trend.

[2]Reference categories.

*Abbreviations: Q, quartile; hPDI, healthful plant-based diet index; uPDI, unhealthful plant-based diet index; BMI, Body Mass Index; HR, hazard ratio; CI, confidence interval.*

**Table *S11*.** Mediation analysis between unhealthful plant-based diet score and chronic kidney disease

| | **Unhealthful Plant-Based Diet Index Score (1-point increments)** | | | | | | | |
|---|---|---|---|---|---|---|---|---|
| | Participants, No. | Total effect **(HR; 95% CI)**[1] | *P*-value | Direct effect **(HR; 95% CI)**[1] | *P*-value | Natural indirect effect **(HR; 95% CI)**[1] | *P*-value | Proportion mediated (Log(NIE) / (Log(NIE)+log(NDE) ) |
| **Potential mediators**[2] | | | | | | | | |
| **Obesity and lipid metabolism** | | | | | | | | |
| BMI | 4,756 | 1.010 [0.995-1.025] | 0.199 | 1.010 [0.994-1.025] | 0.224 | 1.001 [1.000-1.025] | 0.199 | 9% |
| Waist circumference | 4,756 | 1.012 [0.997-1.027] | 0.132 | 1.009 [0.994-1.024] | 0.248 | 1.003 [1.001-1.004] | <0.001 | 25% |
| LDL Cholesterol | 4,744 | 1.009 [0.994-1.025] | 0.234 | 1.009 [0.994-1.025] | 0.241 | 1.000 [0.999-1.000] | 0.452 | NA |
| HDL Cholesterol | 4,361 | 1.009 [0.993-1.025] | 0.281 | 1.008 [0.992-1.024] | 0.337 | 1.001 [1.000-1.002] | 0.060 | 11% |
| Triglycerides | 4,753 | 1.009 [0.994-1.025] | 0.236 | 1.008 [0.993-1.024] | 0.286 | 1.001 [1.000-1.002] | 0.035 | 11% |
| Lipoprotein A | 3,639 | 1.010 [0.993-1.028] | 0.245 | 1.010 [0.993-1.028] | 0.253 | 1.000 [1.000-1.001] | 0.581 | NA |
| Apolipoprotein A | 4,351 | 1.009 [0.993-1.025] | 0.264 | 1.009 [0.993-1.025] | 0.285 | 1.000 [0.999-1.001] | 0.245 | NA |
| **Kidney function** | | | | | | | | |
| eGFR | 4,756 | 1.008 [0.992-1.025] | 0.305 | 1.007 [0.991-1.023] | 0.379 | 1.001 [0.998-1.004] | 0.395 | 13% |
| Cystatin C | 4,754 | 1.009 [0.993-1.025] | 0.282 | 1.001 [0.985-1.017] | 0.894 | 1.008 [1.004-1.011] | <0.001 | 89% |
| Urate | 4,748 | 1.010 [0.994-1.025] | 0.226 | 1.009 [0.994-1.025] | 0.236 | 1.000 [0.999-1.002] | 0.772 | NA |
| Creatinine | 4,756 | 1.009 [0.993-1.025] | 0.256 | 1.005 [0.990-1.021] | 0.508 | 1.004 [1.001-1.007] | 0.009 | 44% |
| Nephrolithiasis | 4,756 | 1.010 [0.994-1.025] | 0.227 | 1.009 [0.994-1.025] | 0.227 | 1.000 [0.999-1.001] | 0.980 | NA |
| **Hypertension** | | | | | | | | |
| Prevalent hypertension | 4,756 | 1.009 [0.992-1.027] | 0.291 | 1.010 [0.995-1.025] | 0.211 | 1.000 [0.992-1.007] | 0.910 | NA |
| Systolic blood pressure | 4,601 | 1.009 [0.994-1.025] | 0.233 | 1.010 [0.994-1.025] | 0.210 | 1.000 [0.999-1.000] | 0.195 | NA |

| | | | | | | | | |
|---|---|---|---|---|---|---|---|---|
| **Inflammatory biomarkers** | | | | | | | | |
| C-reactive protein | 4,746 | 1.010 [0.994-1.025] | 0.218 | 1.010 [0.994-1.025] | 0.235 | 1.000 [0.999-1.001] | 0.503 | NA |

[1]Hazard Ratios with 95% Confidence Intervals (CI) for unhealthful plant-based diet score (1-point increments), adjusted for sex, BMI (excluding when considered as a potential mediator), waist circumference (excluding when considered as a potential mediator), ethnicity, physical activity, smoking status, education, alcohol intake, energy intake, multimorbidity index, Townsend deprivation index, type of diabetes, diabetes medication use, number of completed dietary assessments, and total protein intake; stratified by age (3-year categories) and region.
[2]Potential mediators are modelled on the continuous scale (excluding prevalent nephrolithiasis and prevalent hypertension which are binary).
*Abbreviations: HR, hazard ratios; CI, confidence intervals; BMI, body mass index; eGFR, estimated glomerular filtration rate; NIE, natural indirect effect; NDE, natural direct effect.*

**Table S12.** Hazard ratios (95% confidence intervals) of chronic kidney disease among all individuals with diabetes across sex-specific quartiles (Q) of the healthful plant-based diet index (hPDI) with E-values (n=7,747)

| | HR (95% CI) | P-value | E-value | Upper CI limit |
|---|---|---|---|---|
| **hPDI**[1] | | | | |
| Q1 | 1.00[2] | | | |
| Q2 | 0.89 (0.75-1.06) | 0.20 | 1.39 | 1.00 |
| Q3 | 0.97 (0.81-1.16) | 0.72 | 1.17 | 1.00 |
| Q4 | 0.76 (0.63-0.92) | 0.005 | 1.71 | 1.31 |
| **Sex** | | | | |
| Female | 1.00[2] | | | |
| Male | 1.14 (0.98-1.32) | 0.08 | | |
| **BMI (kg/m$^2$)** | | | | |
| Normal | 1.00[2] | | | |
| Overweight | 1.07 (0.84-1.37) | 0.57 | | |
| Obese | 1.15 (0.87-1.53) | 0.33 | | |
| Missing | 3.84 (1.98-7.45) | <0.001 | | |
| **Waist circumference (cm)** | 1.01 (1.01-1.02) | <0.001 | | |
| **Smoking status** | | | | |
| Never | 1.00[2] | | | |
| Previous | 1.15 (1.01-1.32) | 0.04 | | |
| Current | 1.38 (1.09-1.74) | 0.007 | | |
| Missing | 0.79 (0.32-1.91) | 0.59 | | |
| **Education** | | | | |
| Low | 1.00[2] | | | |
| Medium | 0.93 (0.76-1.13) | 0.46 | | |
| High | 0.85 (0.72-1.00) | 0.05 | | |
| Missing | 1.03 (0.85-1.25) | 0.75 | | |
| **Ethnicity** | | | | |
| White | 1.00[2] | | | |
| Mixed | 1.04 (0.74-1.47) | 0.81 | | |
| Asian | 0.73 (0.49-1.08) | 0.11 | | |
| Black | 0.91 (0.50-1.67) | 0.77 | | |
| Other/Missing | 1.37 (0.90-2.07) | 0.14 | | |
| **Townsend deprivation Index** | | | | |
| Q1 | 1.00[2] | | | |
| Q2 | 0.97 (0.80-1.18) | 0.75 | | |
| Q3 | 0.88 (0.71-1.07) | 0.20 | | |
| Q4 | 0.98 (0.80-1.20) | 0.85 | | |
| Q5 | 1.24 (1.02-1.52) | 0.03 | | |
| **Physical activity (MET-h/wk)** | | | | |
| Q1 | 1.00[2] | | | |
| Q2 | 0.93 (0.77-1.12) | 0.45 | | |
| Q3 | 0.90 (0.74-1.09) | 0.29 | | |
| Q4 | 0.83 (0.68-1.02) | 0.07 | | |
| Q5 | 0.77 (0.63-0.94) | 0.01 | | |
| Missing | 0.90 (0.61-1.32) | 0.59 | | |
| **Energy intake (kJ/day)** | 1.00 (0.99-1.00) | 0.16 | | |
| **Alcohol intake (g/day)** | 0.99 (0.99-1.00) | <0.001 | | |
| **Diabetes medication** | | | | |

| | | |
|---|---|---|
| No | 1.00[2] | |
| Yes | 1.23 (0.93-1.62) | 0.14 |
| Missing | 0.93 (0.69-1.25) | 0.62 |
| **Diabetes type** | | |
| Type 1 | 1.00[2] | |
| Type 2 | 0.80 (0.67-0.95) | 0.009 |
| Unspecified | 0.43 (0.33-0.57) | <0.001 |
| **Protein intake (g/day)** | 1.00 (0.99-1.00) | 0.05 |
| **Number of completed dietary recalls** | 0.99 (0.93-1.04) | 0.64 |

[1]Hazard Ratios with 95% Confidence Intervals (CI) adjusted for sex, education, BMI, waist circumference, ethnicity, physical activity, smoking status, alcohol intake, energy intake, multimorbidity index, townsend depriation index, type of diabetes, diabetes medication use, number of completed dietary assessments, and total protein intake; stratified by age (3-year categories) and region.
[2]Reference categories.
P-trend is for linear trend.
*Abbreviations: Q, quartile; hPDI, healthful plant-based diet index; BMI, Body Mass Index; HR, hazard ratio; CI, confidence interval.*

**Table S13.** Hazard ratios (95% confidence intervals) of chronic kidney disease among all individuals with diabetes across sex-specific quartiles (Q) of the unhealthful plant-based diet index (uPDI) with E-values (n=7,747)

| | HR (95% CI) | P-value | E-value | Upper CI limit |
|---|---|---|---|---|
| **uPDI[1]** | | | | |
| Q1 | 1.00[2] | | | |
| Q2 | 1.20 (1.01-1.43) | 0.04 | 1.53 | 1.09 |
| Q3 | 1.20 (1.00-1.45) | 0.06 | 1.53 | 1.00 |
| Q4 | 1.35 (1.11-1.65) | 0.003 | 1.76 | 1.36 |
| **Sex** | | | | |
| Female | 1.00[2] | | | |
| Male | 1.16 (1.00-1.34) | 0.05 | | |
| **BMI (kg/m$^2$)** | | | | |
| Normal | 1.00[2] | | | |
| Overweight | 1.08 (0.85-1.38) | 0.52 | | |
| Obese | 1.16 (0.88-1.55) | 0.30 | | |
| Missing | 3.95 (2.04-7.68) | <0.001 | | |
| **Waist circumference (cm)** | 1.01 (1.01-1.02) | <0.001 | | |
| **Smoking status** | | | | |
| Never | 1.00[2] | | | |
| Previous | 1.17 (1.02-1.34) | 0.03 | | |
| Current | 1.40 (1.11-1.76) | 0.004 | | |
| Missing | 0.80 (0.33-1.94) | 0.62 | | |
| **Education** | | | | |
| Low | 1.00[2] | | | |
| Medium | 0.93 (0.77-1.13) | 0.49 | | |
| High | 0.85 (0.73-1.01) | 0.06 | | |
| Missing | 1.03 (0.85-1.24) | 0.78 | | |
| **Ethnicity** | | | | |
| White | 1.00[2] | | | |
| Mixed | 1.04 (0.74-1.47) | 0.82 | | |
| Asian | 0.74 (0.50-1.10) | 0.13 | | |
| Black | 0.92 (0.50-1.67) | 0.77 | | |
| Other/Missing | 1.38 (0.91-2.09) | 0.13 | | |
| **Deprivation Index** | | | | |
| Q1 | 1.00[2] | | | |
| Q2 | 0.97 (0.80-1.18) | 0.75 | | |
| Q3 | 0.87 (0.71-1.07) | 0.20 | | |
| Q4 | 0.98 (0.80-1.20) | 0.88 | | |
| Q5 | 1.24 (1.02-1.52) | 0.03 | | |
| **Physical activity (MET-h/wk)** | | | | |
| Q1 | 1.00[2] | | | |
| Q2 | 0.93 (0.77-1.13) | 0.47 | | |
| Q3 | 0.90 (0.74-1.09) | 0.27 | | |
| Q4 | 0.83 (0.68-1.02) | 0.08 | | |
| Q5 | 0.77 (0.63-0.94) | 0.01 | | |
| Missing | 0.91 (0.62-1.34) | 0.63 | | |
| **Energy intake (kJ/day)** | 1.00 (0.99-1.00) | 0.09 | | |
| **Alcohol intake (g/day)** | 0.99 (0.99-1.00) | <0.001 | | |
| **Diabetes medication** | | | | |

| | | |
|---|---|---|
| No | 1.00[2] | |
| Yes | 1.22 (0.92-1.60) | 0.16 |
| Missing | 0.92 (0.69-1.24) | 0.59 |
| **Diabetes type** | | |
| Type 1 | 1.00[2] | |
| Type 2 | 0.79 (0.67-0.94) | 0.007 |
| Unspecified | 0.43 (0.32-0.57) | <0.001 |
| **Protein intake (g/day)** | 1.00 (0.99-1.00) | 0.28 |
| **Number of completed dietary recalls** | 1.01 (0.95-1.07) | 0.75 |

[1]Hazard Ratios with 95% Confidence Intervals (CI) adjusted for sex, education, BMI, waist circumference, ethnicity, physical activity, smoking status, alcohol intake, energy intake, multimorbidity index, townsend depriation index, type of diabetes, diabetes medication use, number of completed dietary assessments, and total protein intake; stratified by age (3-year categories) and region.
[2]Reference categories.
P-trend is for linear trend.
*Abbreviations: Q, quartile; uPDI, unhealthful plant-based diet index; BMI, Body Mass Index; HR, hazard ratio; CI, confidence interval.*

**References**

1. Purcell S, Neale B, Todd-Brown K, Thomas L, Ferreira MA, Bender D, et al. PLINK: a tool set for whole-genome association and population-based linkage analyses. Am J Hum Genet. 2007;81(3):559-75. Epub 2007/08/19. doi: 10.1086/519795. PubMed PMID: 17701901; PubMed Central PMCID: PMCPMC1950838.
2. Chang CC, Chow CC, Tellier LC, Vattikuti S, Purcell SM, Lee JJ. Second-generation PLINK: rising to the challenge of larger and richer datasets. Gigascience. 2015;4:7. Epub 2015/02/28. doi: 10.1186/s13742-015-0047-8. PubMed PMID: 25722852; PubMed Central PMCID: PMCPMC4342193.
3. Bycroft C, Freeman C, Petkova D, Band G, Elliott LT, Sharp K, et al. The UK Biobank resource with deep phenotyping and genomic data. Nature. 2018;562(7726):203-9.
4. Levey AS, Stevens LA, Schmid CH, Zhang Y, Castro III AF, Feldman HI, et al. A new equation to estimate glomerular filtration rate. Annals of internal medicine. 2009;150(9):604-12.
